# Medium-term impacts of the waves of the COVID-19 epidemic on treatments for non-COVID-19 patients in intensive care units: a retrospective cohort study in Japan

**DOI:** 10.1101/2022.02.28.22271604

**Authors:** Shusuke Watanabe, Jung-ho Shin, Takuya Okuno, Tetsuji Morishita, Daisuke Takada, Susumu Kunisawa, Yuichi Imanaka

**Affiliations:** Department of Healthcare Economics and Quality Management, Graduate School of Medicine, Kyoto University, Kyoto City, Kyoto, Japan

## Abstract

**Background:** Maintaining critical care for non-Coronavirus-disease-2019 (non-COVID-19) patients is a key pillar of tackling the impact of the COVID-19 pandemic. This study aimed to reveal the medium-term impacts of the COVID-19 epidemic on case volumes and quality of intensive care for critically ill non-COVID-19 patients.

**Methods:** Administrative data were used to investigate the trends in case volumes of admissions to intensive care units (ICUs) compared with the previous years. Standardized mortality ratios (SMRs) of non-COVID-19 ICU patients were calculated in each wave of the COVID-19 epidemic in Japan.

**Results:** The ratios of new ICU admissions of non-COVID-19 patients to those in the corresponding months before the epidemic: 21% in May 2020, 8% in August 2020, 9% in February 2021, and 14% in May 2021, approximately concurrent with the peaks in COVID-19 infections. The decrease was greatest for new ICU admissions of non-COVID patients receiving mechanical ventilation (MV) on the first day of ICU admission: 26%, 15%, 19%, and 19% in the first, second, third, and fourth waves, respectively. No statistically significant change in SMR was observed in any wave of the epidemic; SMRs were 0.990 (95% confidence interval (CI), 0.962-1.019), 0.979 (95% CI, 0.953-1.006), 0.996 (95% CI, 0.980-1.013), and 0.989 (95% CI, 0.964-1.014), in the first, second, third, and fourth waves of the epidemic, respectively.

**Conclusions:** Compared to the previous years, the number of non-COVID-19 ICU patients continuously decreased over the medium term during the COVID-19 epidemic. The decrease in case volumes was larger in non-COVID-19 ICU patients initially receiving MV than those undergoing other initial treatments. The standardized in-hospital mortality of non-COVID-19 ICU patients did not change in any waves of the epidemic.

**EYWORDS:** Intensive care unit, COVID-19, mechanical ventilation, in-hospital mortality

## INTRODUCTION

Coronavirus disease 2019 (COVID-19) has had wide-ranging impacts on society globally, including many excess deaths[1]. Since most countries have experienced repeated waves of COVID-19[2], it remains a global health concern after first emerging more than two years ago.

Maintaining treatments for non-COVID-19 critically ill patients is recognized as one of the key pillars of tackling the impact of the pandemic. Many countries have applied resource allocation policies for emergency treatments[3] to ensure sufficient capacity to deal with COVID-19 patients, and triaging admissions to intensive care units (ICUs) has been recommended[4]. However, there remain non-COVID-19 patients requiring critical care, and Japanese national policies against COVID-19 stress the importance of concurrently maintaining health systems for non-COVID-19 patients[5].

Since the start of the pandemic, there have been various impacts on critical care for non-COVID-19 patients[6–14]. Reduced ICU utilization during the waves of COVID-19 infections and changes in patient characteristics, such as diagnosis patterns, severity, and admission process, have been reported[6–8]. Furthermore, stresses on medical staff caring for COVID-19 patients have been reported[9, 10], potentially impacting the level of care for non-COVID-19 patients. Some studies examined the impacts on quality of critical care for non-COIVID-19 patients, but conclusion is not in agreement [11–14].

Although the short-term and volume impacts of the COVID-19 pandemic have been reported, impacts on quality over the medium term remain unclear. In addition, while COVID-19 originally emerged in Asia, research in Asia about its impact is scarce. We analyzed a large nationwide claims database in Japan that contained data until approximately one year after the emergence of the COVID-19 pandemic, aiming to clarify the medium-term impacts – impacts during several waves of the COVID-19 – of COVID-19 on critical care for non-COVID-19 patients.

## METHODS

### The COVID-19 epidemic in Japan

Japan has experienced repeating waves of COVID-19[15]. Supplementary Figure 1 shows the daily number of confirmed COVID-19 cases[16]. Referring to the number of newly detected COVID-19 patients, the peaks of the four waves occurred in April 2020, August 2020, January 2021, and May 2021. The Japanese government declared the first state of emergency on April 7^th^, 2020[17]. This declaration was based on recognizing that COVID-19 could have severe impacts on public health and the national economy. Consequently, public health interventions were enabled to suppress infections, such as requesting the public to self-quarantine[17]. The first state of emergency was lifted on May 25^th^, 2020[17], but another state of emergency was declared from January 7^th^, 2021, to March 18^th^, 2021, and from April 23^rd^, 2021 to September 28^th^, 2021[17]. In this study, based on COVID-19 patient trends, April 2020 was set as the start of the COVID-19 epidemic in Japan, and the first, second, third, and fourth waves were set from April to June 2020, July to September 2020, and October 2020 to March 2021, and April 2021 onwards, respectively.

### Data Source

This study utilized data from the Diagnosis Procedure Combination / Per-Diem Payment System (DPC/PDPS) obtained from the Quality Indicator/Improvement Project’s (QIP) database. The QIP database is administered by the Department of Healthcare Economics and Quality Management, Kyoto University School of Public Health. The hospitals participating in the QIP regularly provide the DPC/PDPS data. The participating hospitals primarily provide acute care and represent various sizes and geographical areas. The list of the hospitals participating QIP that have agreed to be made public in advance is offered on the QIP website (http://med-econ.umin.ac.jp/QIP/sanka_byouin.html).

Reimbursement claims in the DPC/PDPS data include the patient’s clinical characteristics and records of clinical services. The clinical characteristics recorded in Form 1 include age, sex, admission date, discharge date, primary diagnosis, and other data. Clinical services are recorded in Files E and F. Major diagnosis categories (MDCs), which are the most medically resource-intensive, are recorded in File D.

### Study Population

From the QIP participating hospitals, those continuously providing DPC/PDPS data from April 2018 to September 2021 were included in our study. Among the patients admitted to these hospitals, ICU admissions from April 2018 to July 2021, aged 18 years or older, were included.

In this study, an ICU was defined as per international standards as the wards which can at least provide oxygen, noninvasive monitoring, and more intensive nursing care than usual beds[18]. Within the general bed reimbursement categories, which are not beds for specified diagnoses (such as stroke) in Japan, three categories meet the ICU criteria as below (the summary description in each category refers to the minimum requirements for reimbursement).

– Specialized-care ICU (sICU): require the most intensive resourcing, including a 1:2 patient-nurse ratio and constant placement of a doctor.
– Emergency-care ICU (eICU): require the facility to deal with emergency patients, a 1:4 patient-nurse ratio, and constant placement of a doctor.
– High care units (HCU): require a 1:5 patient-nurse ratio and constant placement of a doctor.

### Descriptive analysis of ICU admissions for non-COVID-19 patients

The primary outcome of interest was case volumes of non-COVID-19 patients admitted to ICUs. The case volume in each wave of COVID-19 infections was analyzed. To take seasonality into account, a trend of the ratio of the case volume in each month to the case volume in the corresponding month before the pandemic was analyzed. More specifically, monthly case volumes until March 2021 were compared with those in corresponding months one year before, and monthly case volumes from April 2021 to July 2021 were compared with those in corresponding months two years before.

Additionally, the ratios of case volumes of non-COVID-19 patients initially admitted to an sICU (admitted to an sICU on the first day of ICU admissions), of non-COVID-19 ICU patients initially undergoing mechanical ventilation (MV), of non-COVID-19 ICU patients initially administered vasopressors, and of all ICU patients, the ratios of the total case numbers (patients times days) of all ICU patients, and the absolute numbers of COVID-19 ICU patients were described. The classification of COVID-19 patients was based on the diagnoses recorded in the DPC/PDPS data. For diagnoses, the International Statistical Classification of Diseases and Related Health Problems, 10th Revision (ICD-10) was applied, and patients with diagnoses of B34.2 and U07.1, except for suspected diagnoses, were classified as COVID-19 patients.

In addition, the association between hospitals’ acceptance of COVID-19 ICU patients and impacts on non-COVID-19 ICU patient volumes was investigated. Specifically, the included hospitals were classified into three categories described below, which were used to stratify the ratios of case volumes of non-COVID-19 patients to before the epidemic.

– Hospitals that were continuously accepting COVID-19 ICU patients: more than ten patient days of COVID-19 patients in every wave of COVID-19.
– Hospitals that were accepting few COVID-19 ICU patients: less than ten patient days of COVID-19 patients in total in the study period.
– Hospitals that were intermediately accepting COVID-19 ICU patients: hospitals meeting neither of the categories above.

Moreover, another type of hospital classification was employed as a sensitivity analysis as below (hereinafter, referred as the month criteria).

– Hospitals that were continuously accepting COVID-19 ICU patients: at least one COVID-19 ICU patient in every month in the study period.
– Hospitals that were accepting non COVID-19 ICU patients: non COVID-19 ICU patient in the study period.
– Hospitals that were intermediately accepting COVID-19 ICU patients: hospitals meeting neither of the categories above.

To account for regional variation in the COVID-19 epidemic, the restricted analysis above was performed in limited areas where the impact of COVID-19 was considered to be the largest. These areas included Hokkaido, Tochigi, Saitama, Chiba, Tokyo, Kanagawa, Gifu, Aichi, Osaka, Kyoto, Hyogo, Okayama, Hiroshima, Fukuoka, and Okinawa, where the duration of the first or second states of emergency was longer than other areas of Japan[17].

### Changes in initial treatments for non-COVID-19 ICU patients at the start of the epidemic

We investigated changes in the initial treatments for non-COVID-19 ICU patients at the start of the epidemic. Specifically, the proportion of initial treatments that were distinctive for ICU patients, such as extracorporeal membrane oxygenation (ECMO), MV, noninvasive positive pressure ventilation or nasal high-flow therapy (NIPPV/NHF), renal replacement therapy (RRT), and vasopressors[19, 20], were examined. Initial treatments were defined as treatments received on the day of admission to the ICU. The proportion of initial treatments in each wave of the COVID-19 epidemic in Japan were compared with those in the corresponding months in the years before the epidemic — from April 2018 to March 2020.

### Standardized mortality ratios of non-COVID-19 ICU patients during the COVID-19 epidemic

We investigated the change in the standardized mortality ratio of non-COVID-19 ICU patients from the beginning of the epidemic, based on indirect standardization. First, a prediction model for the in-hospital mortality of non-COVID-19 patients was developed based on observations before the epidemic. Second, based on observations after the COVID-19 epidemic began, the ratio of observed in-hospital deaths to expected in-hospital deaths from the prediction model was calculated as a standardized mortality ratio (SMR). The SMR was calculated in each wave of COVID-19 infections, stratified into the categories of hospitals based on acceptance of COVID-19 patients in the ICU.

Although the DPC/PDPS data from the target population of this study did not include risk scores for ICU patients (such as a sequential organ failure assessment (SOFA) score and acute physiology and chronic health evaluation (APACHE) Ⅱ scores), the available data had an acceptable performance with regards to risk adjustment for ICU patients[19, 20]. In this study, sex, age, smoking history, body mass index (BMI), major diagnosis category (MDC), ICU category of initial admission (sICU, eICU, or HCU), initial ICU treatment (ECMO, MV, NIPPV/NHF, RRT, and vasopressors), admission process (post-emergency operation, post elective operation, or medical indication), months of admissions to ICUs, e.g., January or February, were used for predictive analysis.

### Statistical analysis

A chi-square test was performed to compare the proportion of initial treatments of non-COVID-19 ICU patients before and after the epidemic. A statistical significance level of 5% was set.

A logistic regression model was employed to develop the prediction model of in-hospital mortality among non-COVID-19 ICU patients. To account for potential clustering by hospitals, a multilevel model with random intercepts for each hospital was applied[21]. For point estimates and confidence intervals, bootstrap methods were employed[21]. Resampling with replacement from the observed data was repeated 1,000 times, and percentiles (2.5% and 97.5%) from the distribution of SMRs were calculated as the lower and upper limits of the confidence intervals. The fiftieth percentile of the distribution was calculated as the point estimate of the SMRs.

SAS software version 9.4 (SAS Institute Inc., Cary, NC) was used for all statistical analyses; PROC GLIMMIX was used for the multilevel logistic regressions.

## RESULTS

### Patient Characteristics

From April 2018 to September 2021, 242 hospitals continuously provided DPC/PDPS data to the QIP. In total, 529,834 patients meeting the criteria were identified in the study, as shown in Supplementary Figure 2. Among these patients, 10,357 were diagnosed with COVID-19, with the remaining 519,477 patients classified as non-COVID-19 ICU patients. Table 1 shows the characteristics of non-COVID-19 patients admitted to ICUs. Patient demographics and diagnoses were fairly persistent over the start of the COVID-19 epidemic. Within typical treatments, MV (12.7%) and vasopressors (26.4%) were relatively frequent.

**Table 1.**
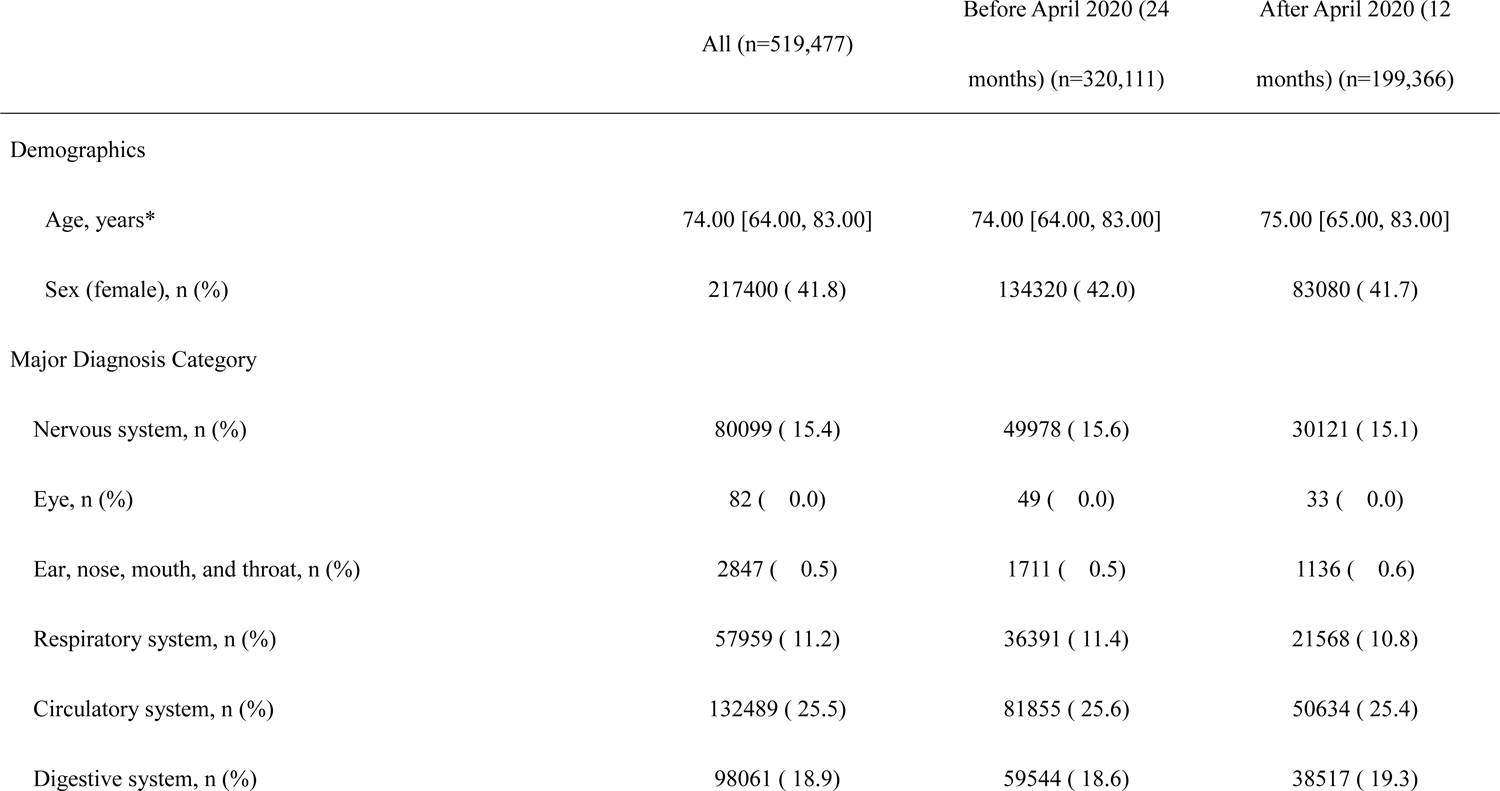
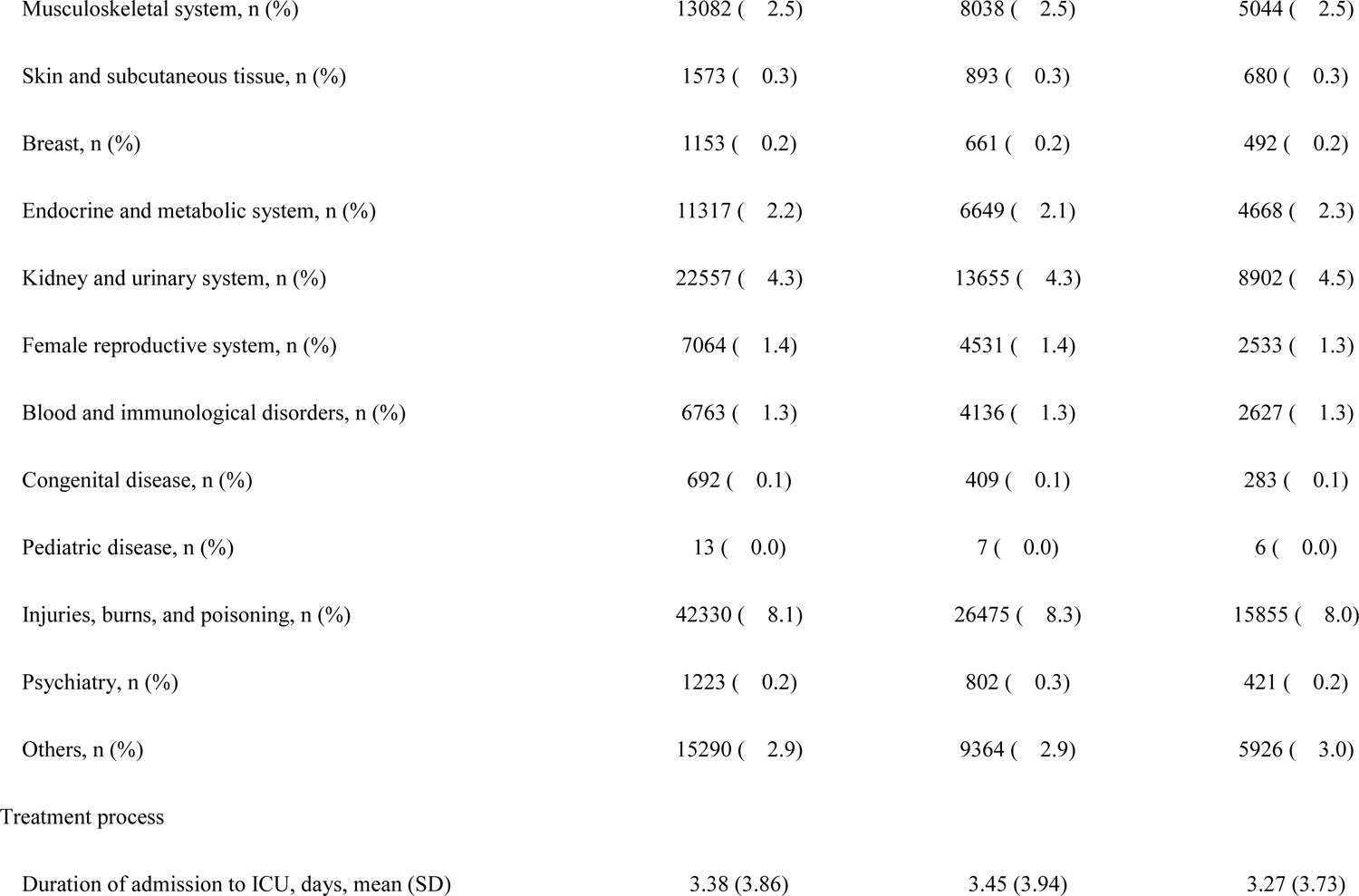
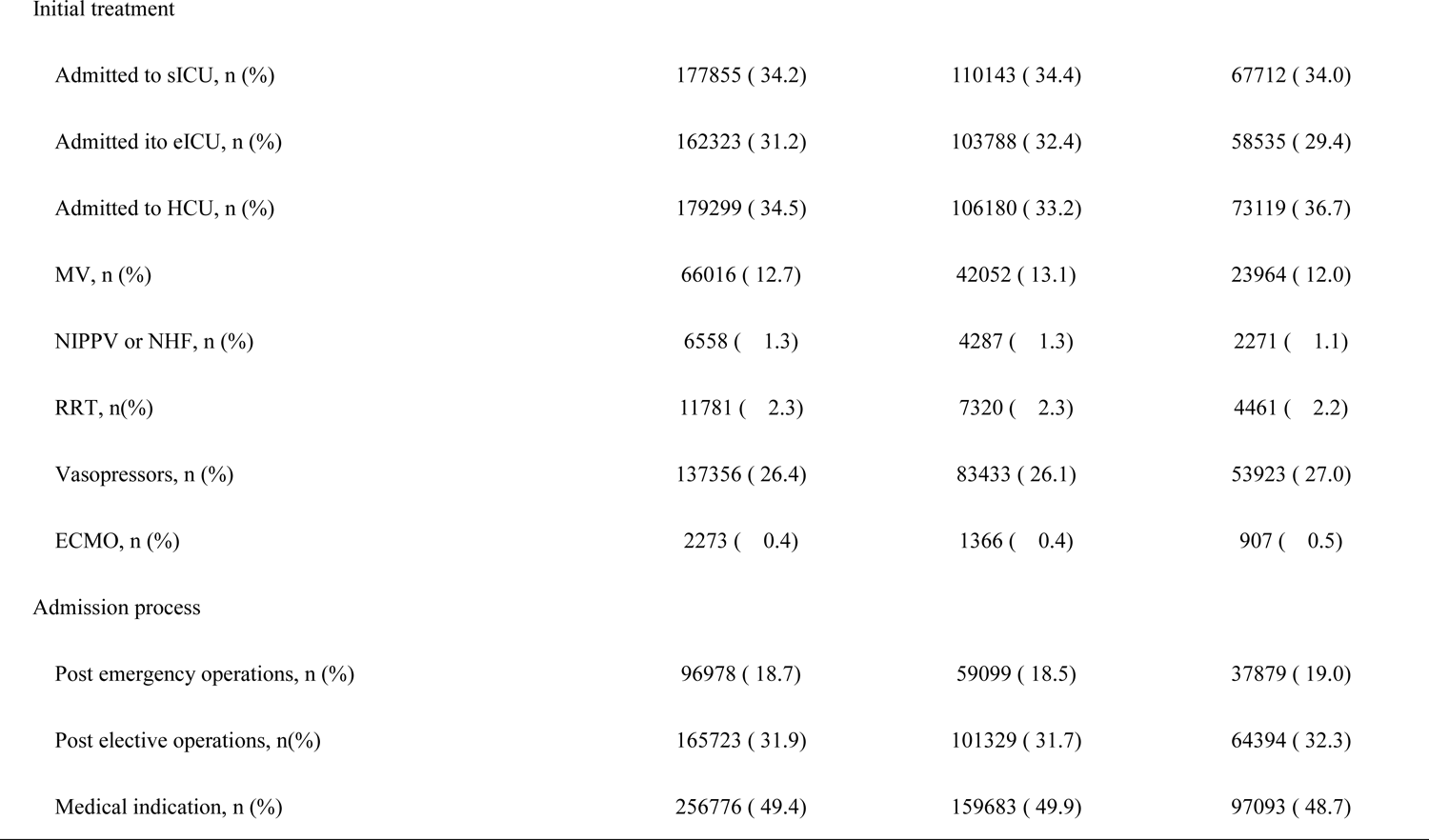
Patient Characteristics

|  | All (n=519,477) | Before April 2020 (24<br>months) (n=320,111) | After April 2020 (12<br>months) (n=199,366) |
| --- | --- | --- | --- |
| Demographics |  |  |  |
| Age, years* | 74.00 [64.00, 83.00] | 74.00 [64.00, 83.00] | 75.00 [65.00, 83.00] |
| Sex (female), n (%) | 217400 ( 41.8) | 134320 ( 42.0) | 83080 ( 41.7) |
| Major Diagnosis Category |  |  |  |
| Nervous system, n (%) | 80099 ( 15.4) | 49978 ( 15.6) | 30121 ( 15.1) |
| Eye, n (%) | 82 ( 0.0) | 49 ( 0.0) | 33 ( 0.0) |
| Ear, nose, mouth, and throat, n (%) | 2847 ( 0.5) | 1711 ( 0.5) | 1136 ( 0.6) |
| Respiratory system, n (%) | 57959 ( 11.2) | 36391 ( 11.4) | 21568 ( 10.8) |
| Circulatory system, n (%) | 132489 ( 25.5) | 81855 ( 25.6) | 50634 ( 25.4) |
| Digestive system, n (%) | 98061 ( 18.9) | 59544 ( 18.6) | 38517 ( 19.3) |
| Musculoskeletal system, n (%) | 13082 ( 2.5) | 8038 ( 2.5) | 5044 ( 2.5) |
| Skin and subcutaneous tissue, n (%) | 1573 ( 0.3) | 893 ( 0.3) | 680 ( 0.3) |
| Breast, n (%) | 1153 ( 0.2) | 661 ( 0.2) | 492 ( 0.2) |
| Endocrine and metabolic system, n (%) | 11317 ( 2.2) | 6649 ( 2.1) | 4668 ( 2.3) |
| Kidney and urinary system, n (%) | 22557 ( 4.3) | 13655 ( 4.3) | 8902 ( 4.5) |
| Female reproductive system, n (%) | 7064 ( 1.4) | 4531 ( 1.4) | 2533 ( 1.3) |
| Blood and immunological disorders, n (%) | 6763 ( 1.3) | 4136 ( 1.3) | 2627 ( 1.3) |
| Congenital disease, n (%) | 692 ( 0.1) | 409 ( 0.1) | 283 ( 0.1) |
| Pediatric disease, n (%) | 13 ( 0.0) | 7 ( 0.0) | 6 ( 0.0) |
| Injuries, burns, and poisoning, n (%) | 42330 ( 8.1) | 26475 ( 8.3) | 15855 ( 8.0) |
| Psychiatry, n (%) | 1223 ( 0.2) | 802 ( 0.3) | 421 ( 0.2) |
| Others, n (%) | 15290 ( 2.9) | 9364 ( 2.9) | 5926 ( 3.0) |
| Treatment process |  |  |  |
| Duration of admission to ICU, days, mean (SD) | 3.38 (3.86) | 3.45 (3.94) | 3.27 (3.73) |
| Initial treatment |  |  |  |
| Admitted to sICU, n (%) | 177855 ( 34.2) | 110143 ( 34.4) | 67712 ( 34.0) |
| Admitted ito eICU, n (%) | 162323 ( 31.2) | 103788 ( 32.4) | 58535 ( 29.4) |
| Admitted to HCU, n (%) | 179299 ( 34.5) | 106180 ( 33.2) | 73119 ( 36.7) |
| MV, n (%) | 66016 ( 12.7) | 42052 ( 13.1) | 23964 ( 12.0) |
| NIPPV or NHF, n (%) | 6558 ( 1.3) | 4287 ( 1.3) | 2271 ( 1.1) |
| RRT, n(%) | 11781 ( 2.3) | 7320 ( 2.3) | 4461 ( 2.2) |
| Vasopressors, n (%) | 137356 ( 26.4) | 83433 ( 26.1) | 53923 ( 27.0) |
| ECMO, n (%) | 2273 ( 0.4) | 1366 ( 0.4) | 907 ( 0.5) |
| Admission process |  |  |  |
| Post emergency operations, n (%) | 96978 ( 18.7) | 59099 ( 18.5) | 37879 ( 19.0) |
| Post elective operations, n(%) | 165723 ( 31.9) | 101329 ( 31.7) | 64394 ( 32.3) |
| Medical indication, n (%) | 256776 ( 49.4) | 159683 ( 49.9) | 97093 ( 48.7) |

### Trends in case volumes of admissions to ICUs

Figure 1 and Supplementary Table 1 show the trends in the ratios of case volumes of admissions to ICUs in each month to before the epidemic. The trends in case numbers of COVID-19 patients admitted to ICUs were similar to the national trends in confirmed COVID-19 cases in Japan, shown in Supplementary Figure 1; the first peak was in April 2020, the second peak was in August 2020, the third peak was in January 2021, and the fourth peak was in May 2021. The third and fourth peaks were larger in size.

**Figure 1.**
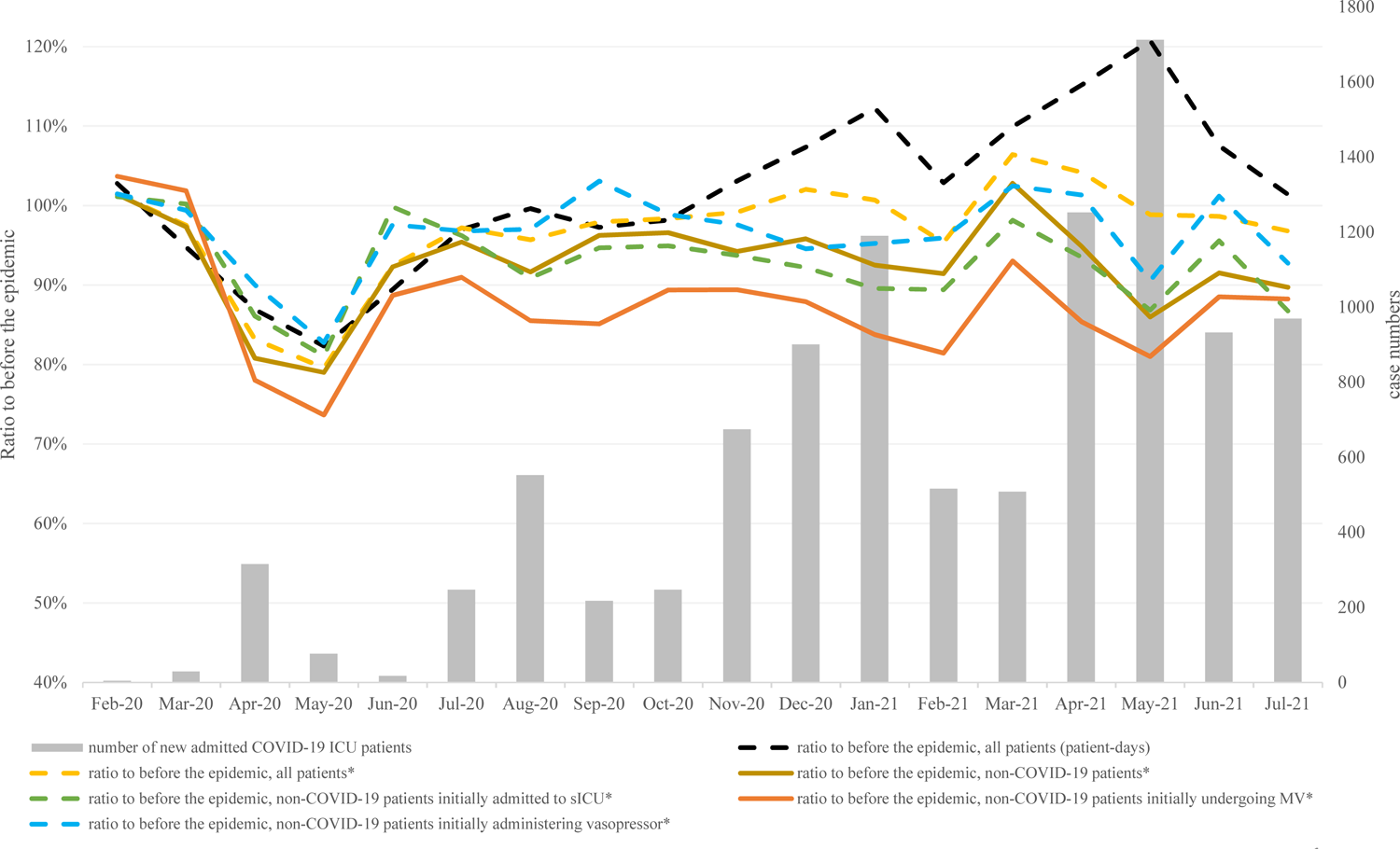
The trends in the ratios of case volumes of admissions to ICUs in each month to the same month before the epidemic

The ratios of new ICU admissions of non-COVID-19 patients declined around the same time as the four peaks in the number of COVID-19 patient admissions to ICUs: a 21% decrease in May 2020, an 8% decrease in August 2020, an 9% decrease in February 2021, and a 14% decrease in May 2021. Similarly, the ratios of new ICU admissions of non-COVID-19 patients initially receiving MV decreased in the same months, but to a greater degree: a 26% decrease in May 2020, a 15% decrease in August 2020, an 19% decrease in February 2021, and 19% decrease in May 2021.

Figure 2 and Supplementary Table 2 show the ratios of non-COVID-19 ICU patients, stratified by hospitals’ acceptance of COVID-19 ICU patients. The decreases in case volumes of non-COVID-19 ICU patients were larger in hospitals continuously accepting COVID-19 ICU patients compared with other categories of hospitals. Supplementary Figure 3 and Supplementary Table 3 show the same data, limited to the areas where the impact of the COVID-19 epidemic was largest. A similar difference in case volumes of non-COVID-19 patients can be observed between hospital categories. Supplementary Figure 4 and Supplementary Table 4 show the same data based on the hospital classification of the month criteria. The same trend of the difference of the impact by the degree of acceptance of COVID-19 ICU patients was observed.

**Figure 2.**
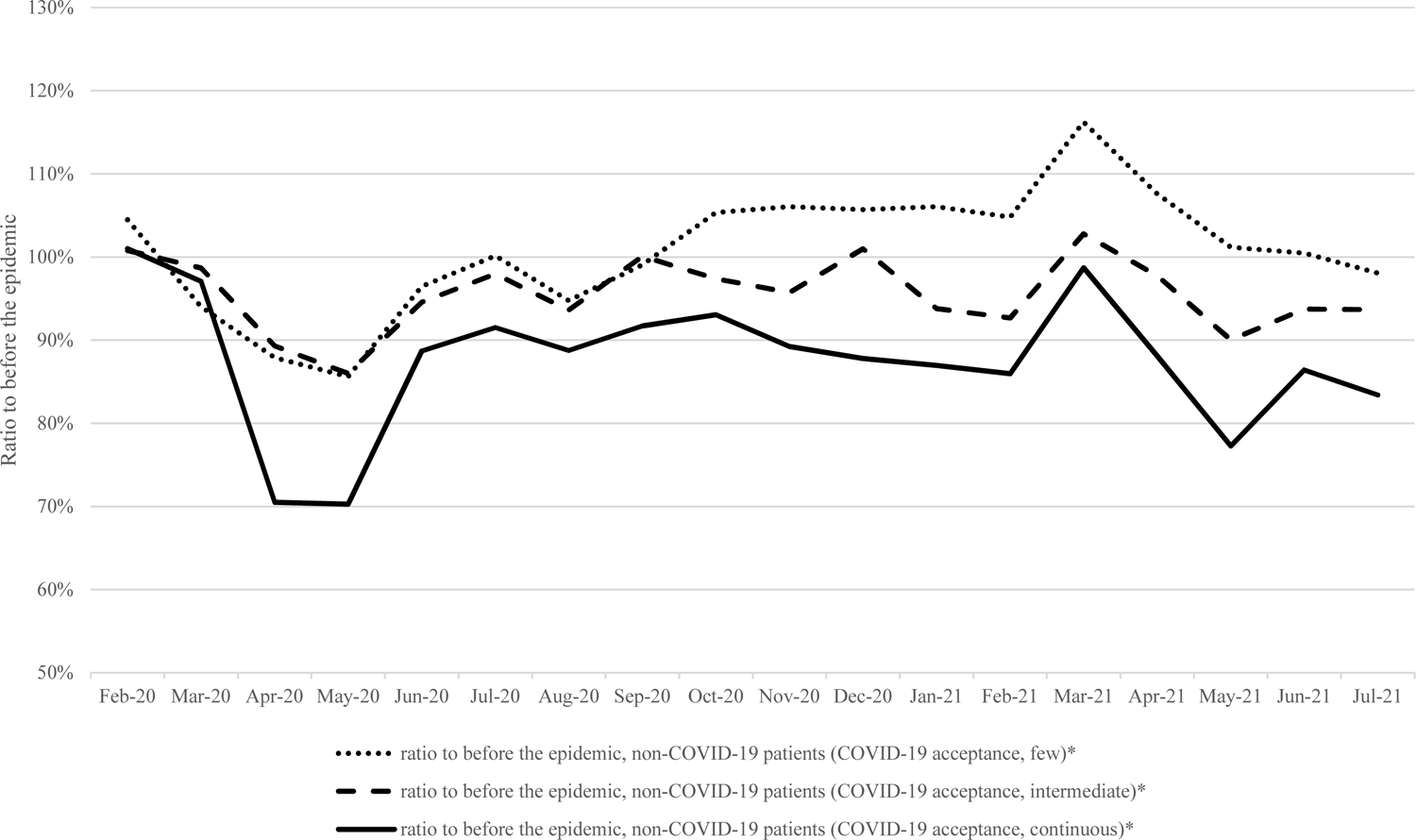
The trends in the ratios of case volumes of admissions of non-COVID-19 ICU patients in each month to the same month before the epidemic, stratified by hospitals

### Changes in initial treatments for non-COVID-19 ICU patients at the start of the COVID-19 epidemic

Figure 3 (MV and vasopressor) and Supplementary Figure 5 (NIPPV/NHF, RRT, and ECMO) show the changes in initial treatments for non-COVID-19 ICU patients. The proportion of patients receiving MV decreased significantly in all waves of the epidemic: 12.7% to 12.2% (p = .0301) in the first wave, 12.0% to 10.8% (p < .0001) in the second wave, 13.9% to 12.7% (p < .0001) in the third wave, and 12.4% to 11.8% (p = .0006) in the fourth wave. On the other hand, the proportion of patients administered vasopressors increased significantly in all waves of the epidemic: 25.8% to 27.5% (p < .0001) in the first wave, 25.1% to 26.1% (p = .0032) in the second wave, 26.6% to 27.1% (p = .0168) in the third wave, and 25.7% to 27.2% (p < .0001) in the fourth wave.

**Figure 3.**
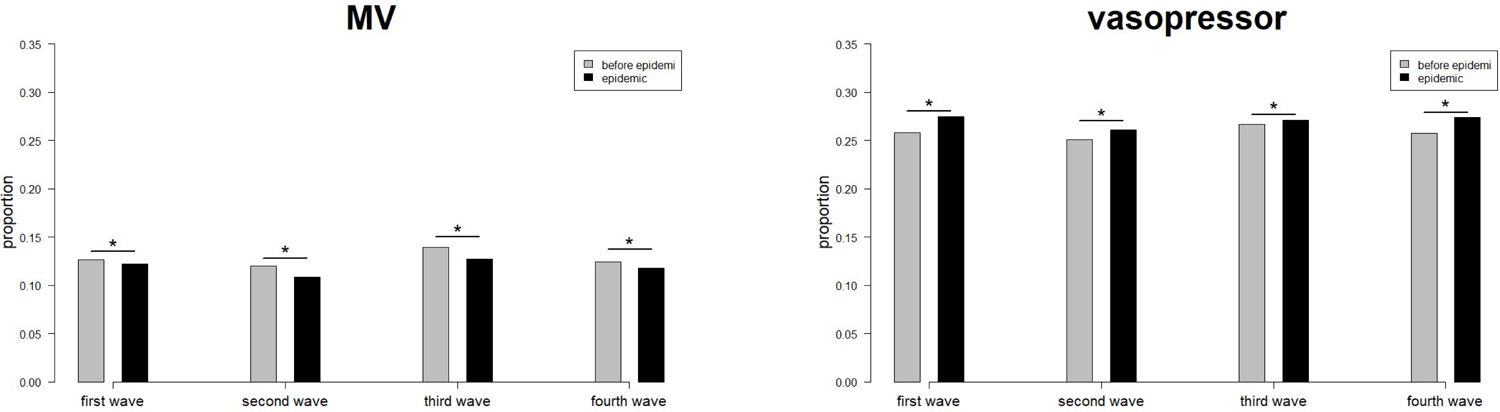
Changes in the proportion of initial treatments received by non-COVID-19 ICU patients (MV and vasopressors)

### Standardized mortality ratios of non-COVID-19 ICU patients during the COVID-19 epidemic

The c-statistic of the prediction model for in-hospital mortality of ICU patients was 0.8776 (95% confidence interval, 0.8756-0.87895). The odds ratios of the model’s predictors are shown in Supplementary Table 5.

Table 2 shows the standardized mortality ratios of ICU patients admitted in each wave of the epidemic. The confidence intervals of SMRs included one in all waves of the epidemic. Supplementary Table 6 shows the SMRs stratified into the hospital categories. There was no statistically significant increase in SMRs, in any of the categories of hospitals, and in any of the waves.

**Table 2.**
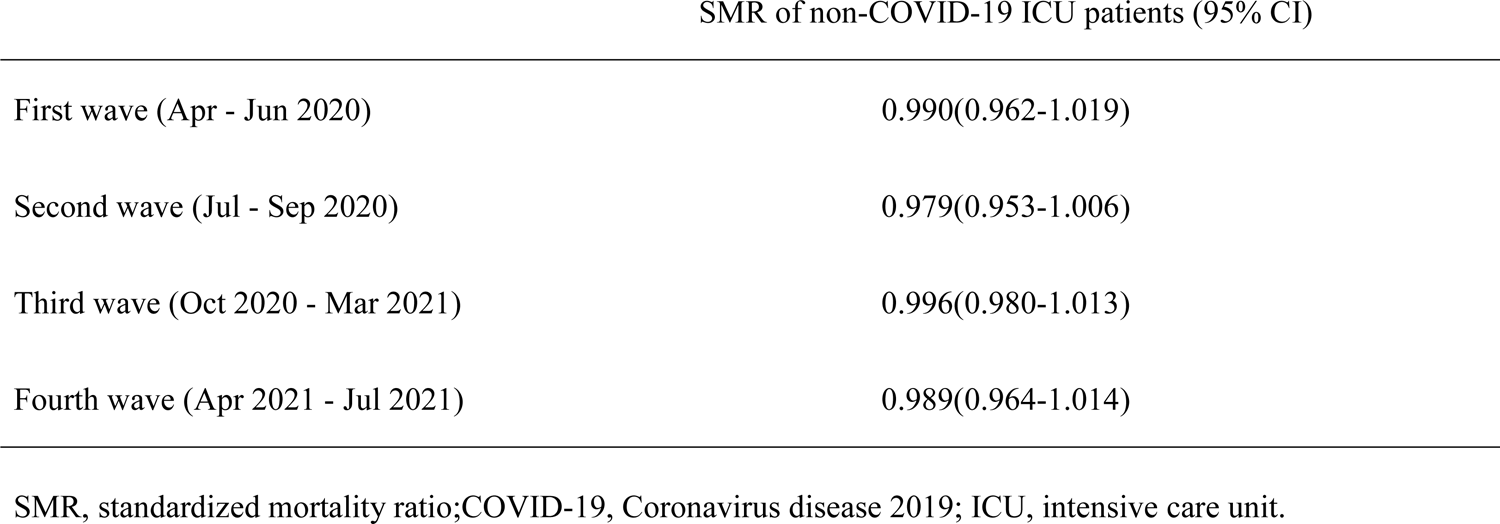
Standardized mortality ratios of non-COVID-19 ICU patients in each wave of the epidemic

## DISCUSSION

Our study investigated a large claims database in Japan and revealed the medium-term impact of waves of the COVID-19 epidemic on the case volumes, initial treatments, and in-hospital mortality ratios of non-COVID-19 patients admitted to ICUs.

Descriptive analysis revealed that the waves of the COVID-19 epidemic negatively impacted the volume of non-COVID-19 ICU admissions. A previous study [6] suggested that the reduced volume of ICU patients was due to patients’ hesitancy to visit hospitals. In fact, a reduction in case volumes has been observed in many areas, including acute coronary syndrome[22], pneumonia[23], and surgeries[24]. However, the reasons behind the reduction in admissions have not been clarified. In addition to patient hesitancy, another potential mechanism of reduced case volumes is the postponement of non-emergency treatments or tests to reserve capacity for critically ill COVID-19 patients. Postponing non-essential procedures was implemented in various countries around the world[3, 4], and the Japanese government also requested health care providers to postpone non-emergency medical procedures during waves of COVID-19 infections[25]. We observed that the decrease in non-COVID-19 ICU patient admissions was greater for hospitals continuously accepting COVID-19 ICU patients, results that are consistent with the latter mechanism of postponement of non-emergency procedures.

This study observed that the impact on case volumes was largest in the first wave compared to subsequent waves. In other countries, decreased patient volumes during subsequent waves of COVID-19 have been reported, but comparisons with the impact of the first wave are not in agreement with the present study[26, 27]. One potential explanation for the smaller impact in subsequent waves is the more organized management of ICU beds. For instance, during subsequent waves, health professionals might take advantage of lessons learned during the first wave. Another possible explanation is less hesitancy to visit hospitals during subsequent waves. Residents may have gradually become accustomed to the COVID-19 epidemic. In fact, reduced mobility in public spaces in Japan was reported to be smaller in subsequent waves than in the first wave[28].

The impact of the COVID-19 epidemic on volumes of non-COVID-19 ICU patients receiving MV was larger than for other treatments. In addition, the proportion of patients receiving MV decreased in all waves of the epidemic. Since the treatment of COVID-19 patients is considered to exhaust the capacity to undertake MV[4], MVs are supposed to have been suppressed more than other treatments. However, changes in patient volumes have been reported in various areas, including acute coronary syndrome[22], pneumonia[23], and surgeries[24]; the influence of these changes cannot be denied. In addition, volumes themselves of respiratory infection diseases were suggested to decrease due to infection prevention measures employed by residents during the epidemic of the COVID-19[23, 29].

Our finding that the confidence intervals of SMRs included one in all waves of the epidemic implies that treatment quality for non-COVID-19 ICU patients was maintained in Japan. Global evidence has been inconsistent about the quality of intensive care during the COVID-19 pandemic[7,8,12–14]. Many factors influence the epidemic’s impact on the quality of intensive care. Confirmed COVID-19 patient numbers were smaller in Japan than in other high-income countries[30], which might explain our findings.

This study has several limitations. First, although the characteristics of the data-providing hospitals were varied, the data collection relied on the voluntary participation of the hospitals. This may introduce selection bias and limit the generalizability of our findings. Second, the DPC/PDPS data of the study population did not include risk scores of ICU patient severity, such as SOFA or APACHE Ⅱ scores. Although the prediction performance was good in our study, the risk was adjusted in different ways compared to other studies[7, 8]. Third, data about the demand for treatments is not available. As mentioned in the previous section, it is difficult to distinguish between the suppression of required treatments and a decline in the demand for treatments. Further research is warranted, including an investigation of the trend in disease volumes in the general population.

## CONCLUSION

We revealed that the number of non-COVID-19 ICU patients continuously decreased over the medium term during the COVID-19 epidemic, compared to the previous year. The decrease in case volumes was larger among non-COVID-19 ICU patients initially receiving MV than those undergoing other initial treatments. The standardized in-hospital mortality of non-COVID-19 ICU patients did not change in any waves of the epidemic.

## Data Availability

Data cannot be shared for ethical/privacy reasons.

## DECRALATION

### ETHICS APPROVAL AND CONSENT TO PARTICIPATE

This study was conducted in accordance with the principles of the Declaration of Helsinki and the study was approved by the Ethics Committee, Kyoto University Graduate School and Faculty of Medicine (approval number: R0135) with a waiver of informed consent prior to data collection.

## CONSENT FOR PUBLICATION

Not applicable

## AVAILABILITY OF DATA AND MATERIALS

Data cannot be shared for ethical/privacy reasons.

## AUTHORS’ CONTRIBUTIONS

S.W., J.S., T.O., T.M., D.T., S.K., and Y.I. were involved in the conceptualization of the study and the design of the methodology. J.S. and S.K. were in charge of the data curation. S.W. and J.S. undertook the formal analysis. S.W. wrote the original draft of the manuscript with input from all authors. All authors critically revised the report, commented on drafts of the manuscript, and approved the final report. Y.I. was in charge of the administration and supervision of the project administration.

## Acknowledgements

We thank all the staff members and all the participating acute care hospitals.

---

**Supplementary Figure 1.**
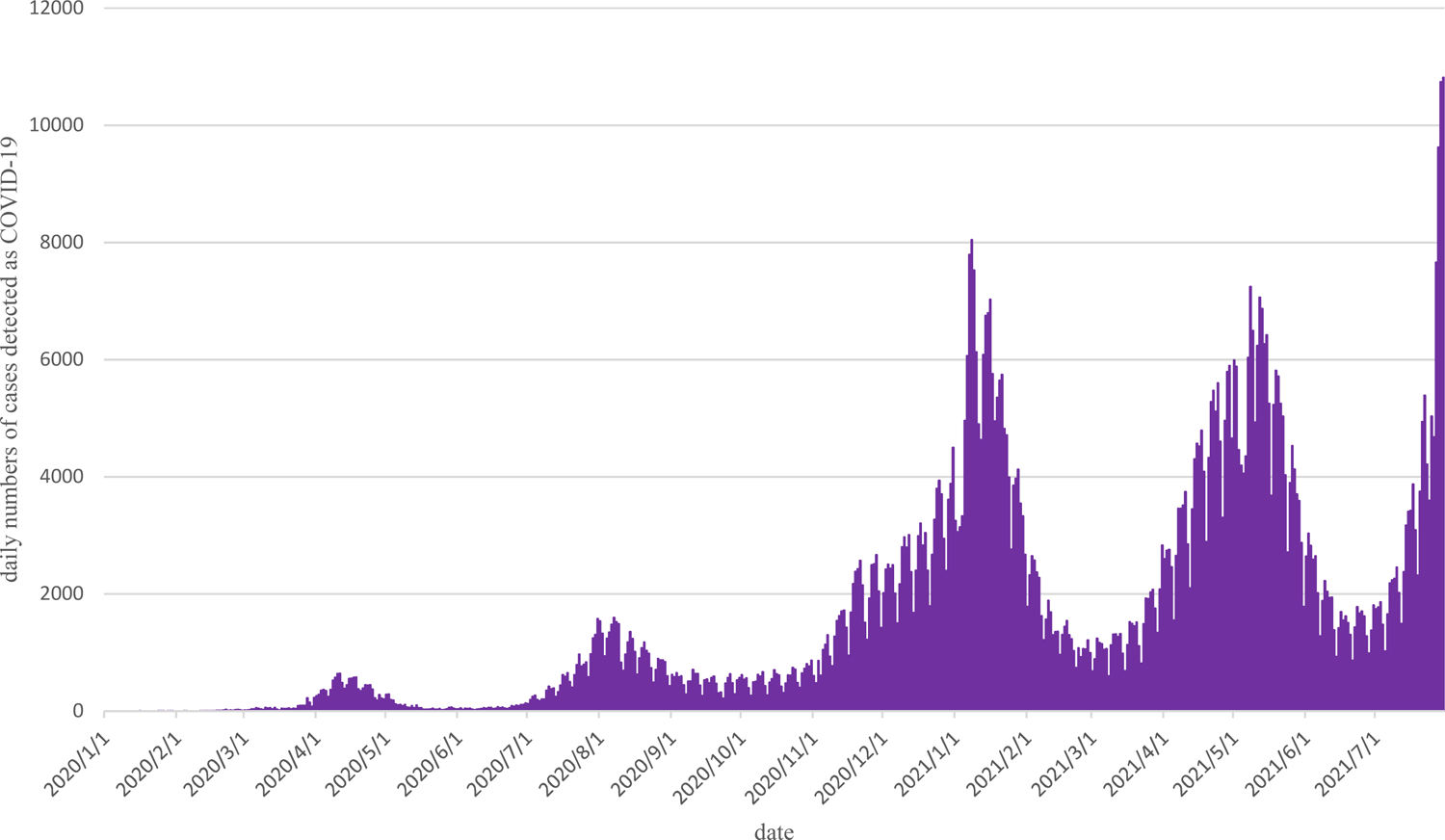
The trend in daily COVID-19 case numbers in Japan

**Supplementary Figure 2.**
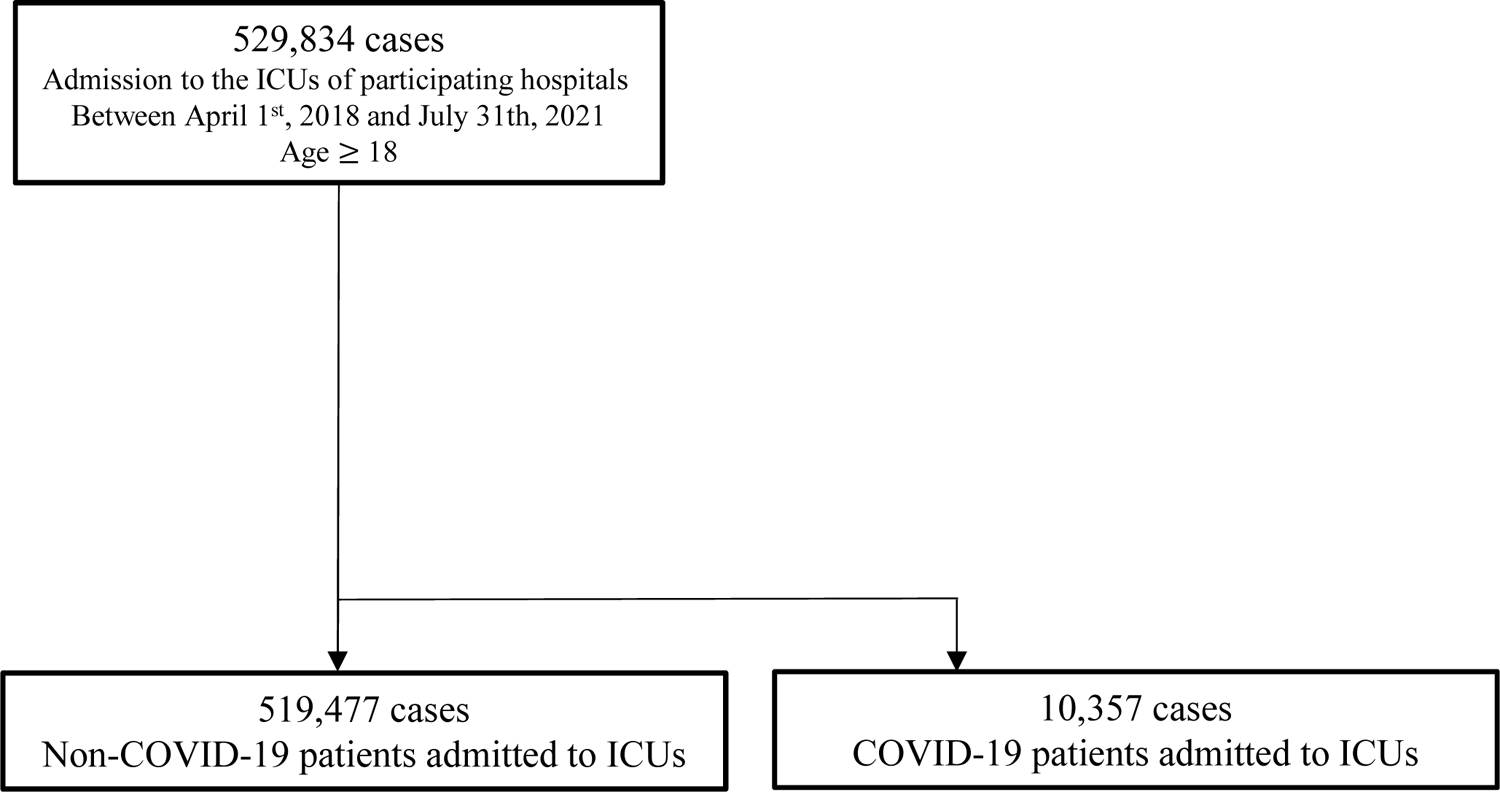
Flowchart of case classifications in this study

**Supplementary Figure 3.**
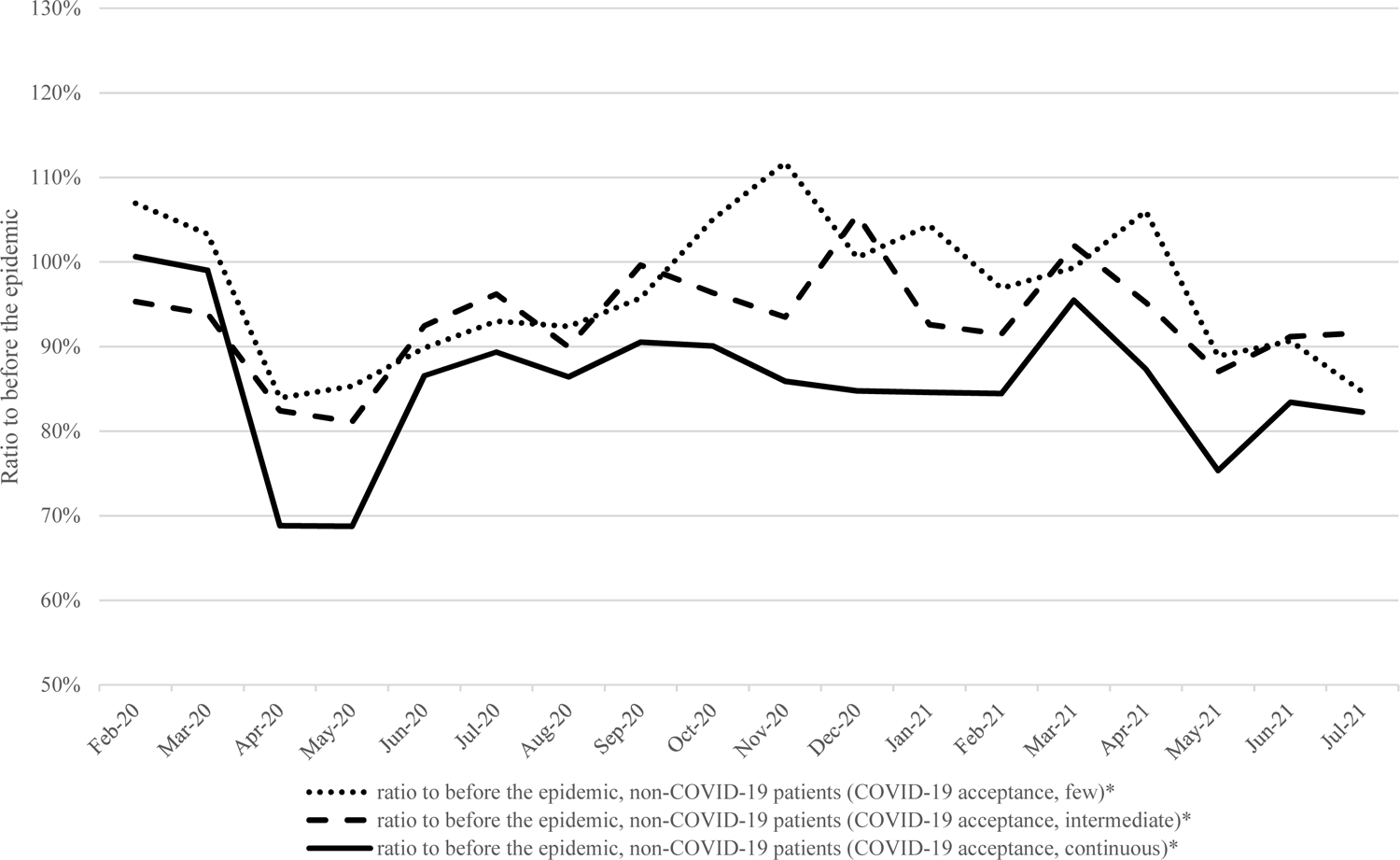
The trend in the ratios of case volumes of admissions of non-COVID-19 ICU patients in each month to the same month before the epidemic, stratified by hospitals, in the prefectures with proactive COVID-19 policies

**Supplementary Figure 4.**
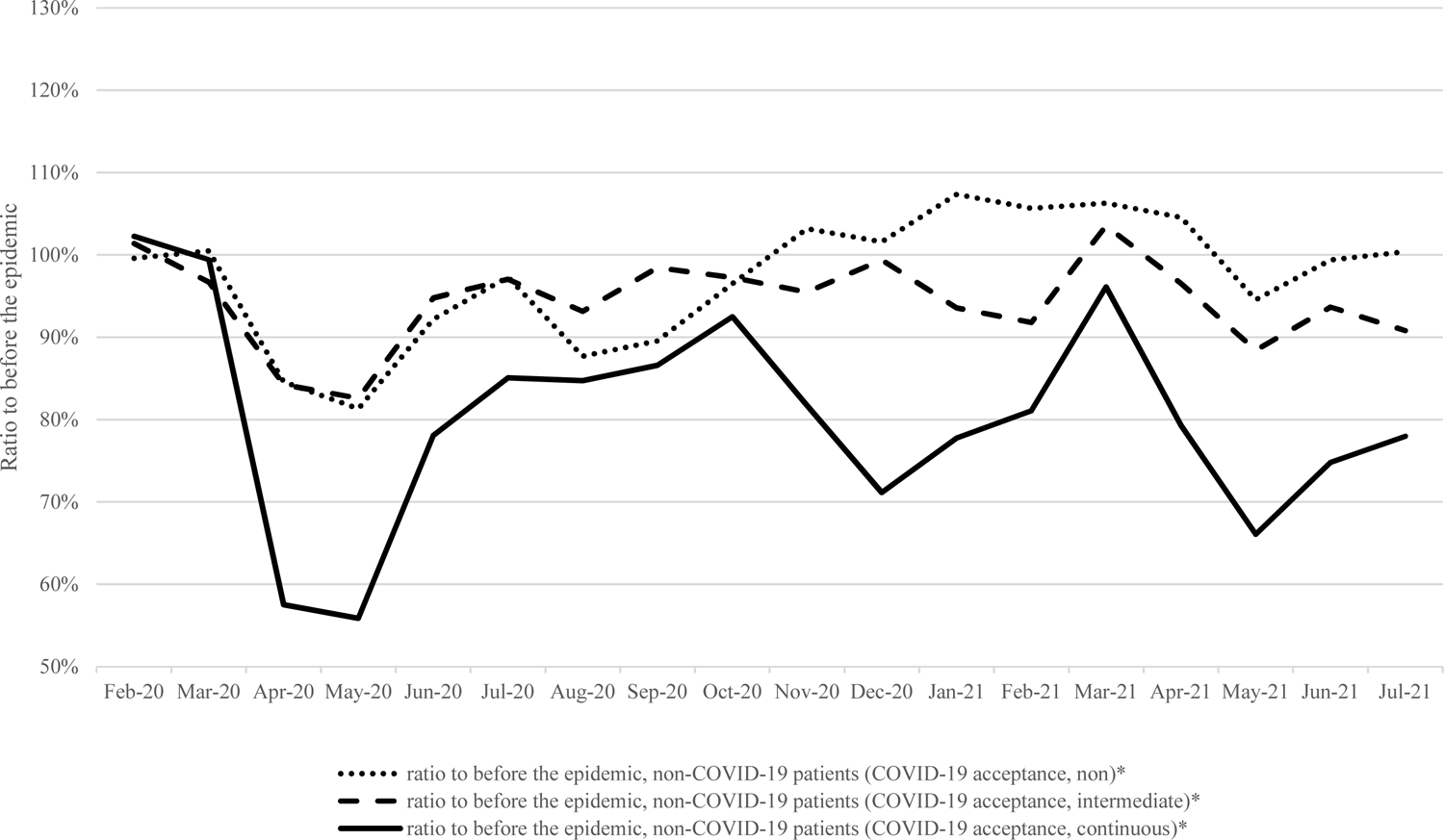
The trend in the ratios of case volumes of admissions of non-COVID-19 ICU patients in each month to the same month before the epidemic, stratified by hospitals (classified by the month criteria)

**Supplementary Figure 5.**
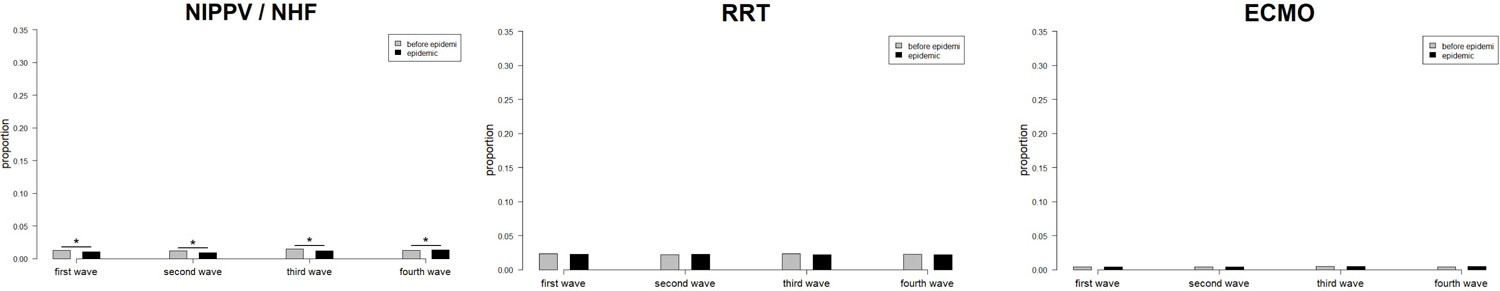
Changes in the proportion of initial treatments for non-COVID-19 ICU patients (NIPPV/NHF, RRT, and ECMO)

**Supplementary Table 1.**
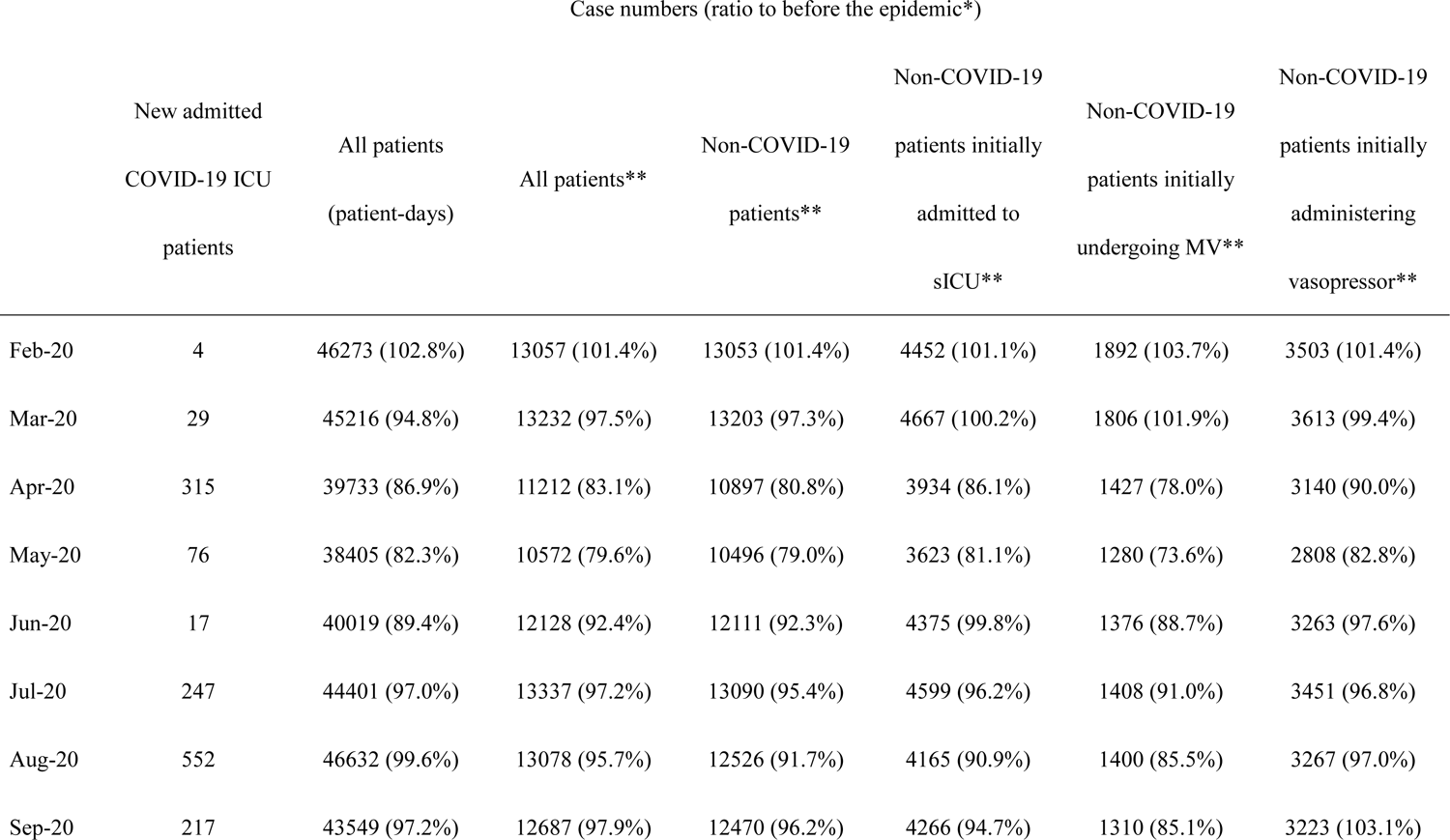
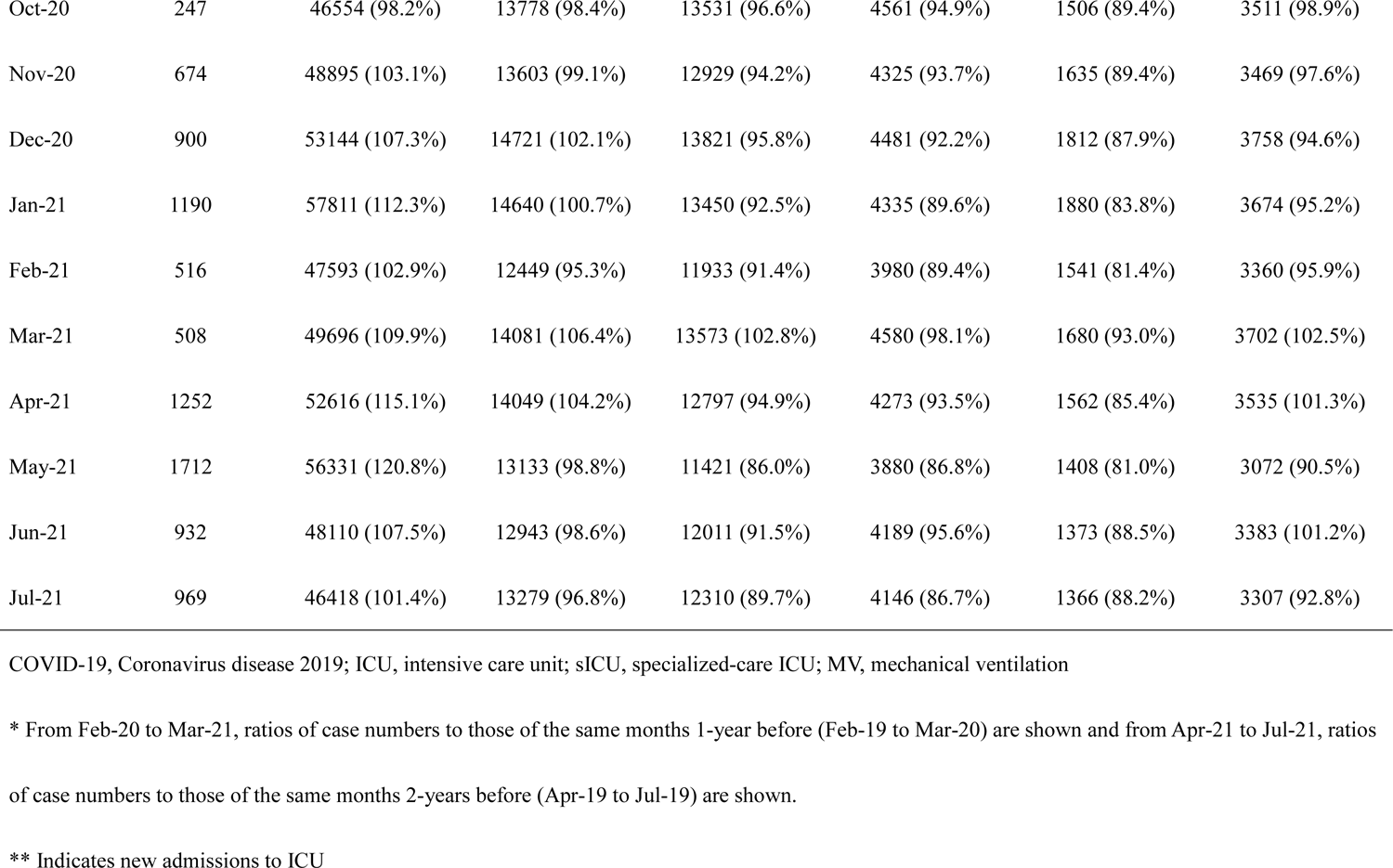
Trends in the ratios of case volumes of admissions to ICUs in each month to the corresponding month before the epidemic Case numbers (ratio to before the epidemic*)

**Supplementary Table 2.**
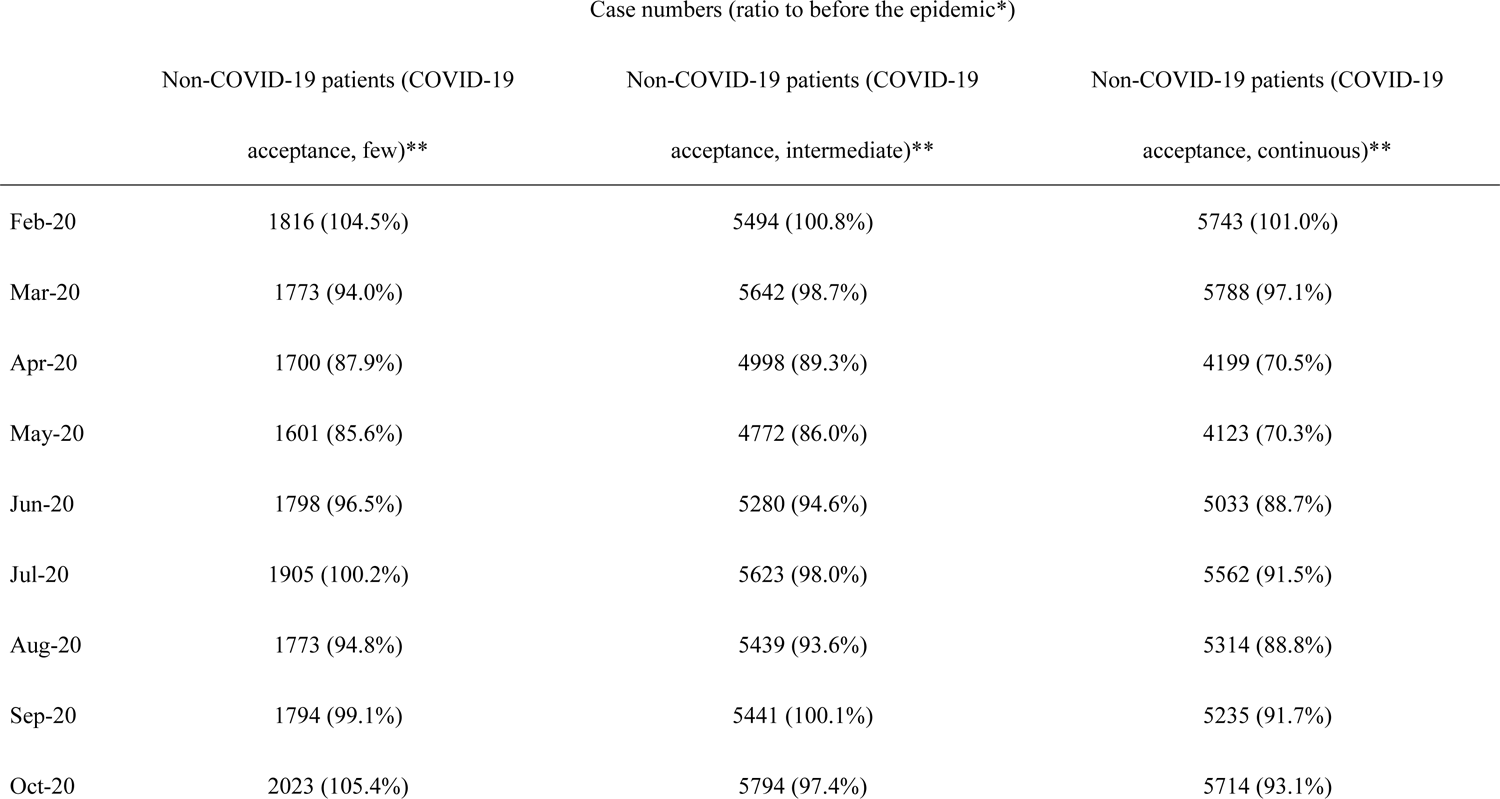
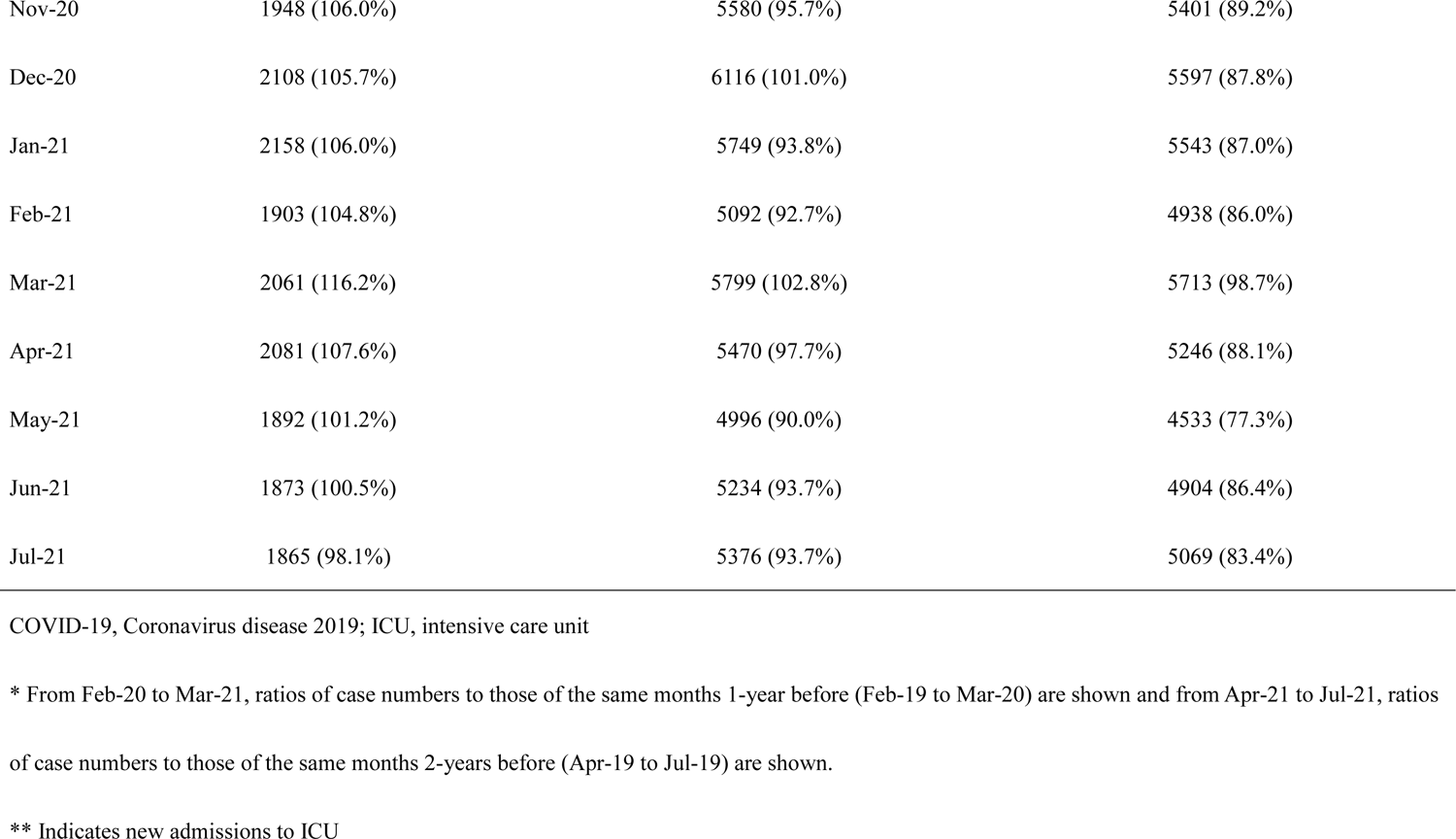
Trends in the ratios of case volumes of non-COVID-19 patient admissions to ICUs in each month to the same month in the previous year, stratified by hospitals

**Supplementary Table 3.** Trends in the ratios of case volumes of non-COVID-19 patient admissions to ICUs in each month to the same month in the previous year, stratified by hospitals, in the prefectures with proactive COVID-19 policies

|  | Case numbers (ratio to before the epidemic*) |  |  |
| --- | --- | --- | --- |
|  | Non-COVID-19 patients (COVID-19 acceptance, few)** | Non-COVID-19 patients (COVID-19 acceptance, intermediate)** | Non-COVID-19 patients (COVID-19 acceptance, continuous)** |
| Feb-20 | 708 (106.9%) | 3025 (95.3%) | 4594 (100.6%) |
| Mar-20 | 723 (103.3%) | 3109 (93.9%) | 4706 (99.0%) |
| Apr-20 | 601 (83.9%) | 2627 (82.4%) | 3299 (68.8%) |
| May-20 | 621 (85.3%) | 2566 (81.1%) | 3252 (68.7%) |
| Jun-20 | 653 (89.8%) | 2936 (92.5%) | 4001 (86.6%) |
| Jul-20 | 691 (93.0%) | 3196 (96.2%) | 4412 (89.3%) |
| Aug-20 | 704 (92.4%) | 3071 (90.0%) | 4185 (86.4%) |
| Sep-20 | 675 (95.7%) | 3103 (99.6%) | 4151 (90.5%) |
| Oct-20 | 768 (105.1%) | 3203 (96.4%) | 4480 (90.1%) |
| Nov-20 | 761 (111.7%) | 3031 (93.5%) | 4210 (85.9%) |
| Dec-20 | 756 (100.5%) | 3486 (105.6%) | 4359 (84.8%) |
| Jan-21 | 817 (104.3%) | 3150 (92.6%) | 4336 (84.6%) |
| Feb-21 | 686 (96.9%) | 2768 (91.5%) | 3880 (84.5%) |
| Mar-21 | 718 (99.3%) | 3170 (102.0%) | 4494 (95.5%) |
| Apr-21 | 759 (106.0%) | 3035 (95.2%) | 4186 (87.3%) |
| May-21 | 647 (88.9%) | 2754 (87.0%) | 3565 (75.4%) |
| Jun-21 | 659 (90.6%) | 2895 (91.2%) | 3855 (83.4%) |
| Jul-21 | 629 (84.7%) | 3044 (91.6%) | 4061 (82.2%) |

COVID-19, Coronavirus disease 2019; ICU, intensive care unit
\* From Feb-20 to Mar-21, ratios of case numbers to those of the same months 1-year before (Feb-19 to Mar-20) are shown and from Apr-21 to Jul-21, ratios of case numbers to those of the same months 2-years before (Apr-19 to Jul-19) are shown.
\*\* Indicates new admissions to ICU

**Supplementary Table 4.** Trends in the ratios of case volumes of non-COVID-19 patient admissions to ICUs in each month to the same month in the previous year, stratified by hospitals (classified by the month criteria)

|  | Case numbers (ratio to before the epidemic*) |  |  |
| --- | --- | --- | --- |
|  | Non-COVID-19 patients (COVID-19 acceptance, non)** | Non-COVID-19 patients (COVID-19 acceptance, intermediate)** | Non-COVID-19 patients (COVID-19 acceptance, continuous)** |
| Feb-20 | 968 (99.6%) | 10366 (101.4%) | 1719 (102.3%) |
| Mar-20 | 1000 (100.5%) | 10500 (96.7%) | 1703 (99.4%) |
| Apr-20 | 874 (84.6%) | 9010 (84.3%) | 1013 (57.5%) |
| May-20 | 823 (81.3%) | 8698 (82.6%) | 975 (55.9%) |
| Jun-20 | 930 (92.2%) | 9773 (94.8%) | 1408 (78.1%) |
| Jul-20 | 980 (97.3%) | 10484 (97.1%) | 1626 (85.1%) |
| Aug-20 | 883 (87.7%) | 10151 (93.1%) | 1492 (84.7%) |
| Sep-20 | 858 (89.6%) | 10150 (98.4%) | 1462 (86.6%) |
| Oct-20 | 992 (96.5%) | 10860 (97.3%) | 1679 (92.5%) |
| Nov-20 | 1001 (103.2%) | 10450 (95.5%) | 1478 (81.7%) |
| Dec-20 | 1072 (101.6%) | 11385 (99.4%) | 1364 (71.2%) |
| Jan-21 | 1123 (107.4%) | 10858 (93.6%) | 1469 (77.8%) |
| Feb-21 | 1023 (105.7%) | 9516 (91.8%) | 1394 (81.1%) |
| Mar-21 | 1063 (106.3%) | 10873 (103.6%) | 1637 (96.1%) |
| Apr-21 | 1080 (104.5%) | 10321 (96.5%) | 1396 (79.3%) |
| May-21 | 957 (94.6%) | 9311 (88.4%) | 1153 (66.1%) |
| Jun-21 | 1003 (99.4%) | 9659 (93.7%) | 1349 (74.8%) |
| Jul-21 | 1011 (100.4%) | 9809 (90.8%) | 1490 (78.0%) |

COVID-19, Coronavirus disease 2019; ICU, intensive care unit
\* From Feb-20 to Mar-21, ratios of case numbers to those of the same months 1-year before (Feb-19 to Mar-20) are shown and from Apr-21 to Jul-21, ratios of case numbers to those of the same months 2-years before (Apr-19 to Jul-19) are shown.
\*\* Indicates new admissions to ICU

**Supplementary Table 5.**
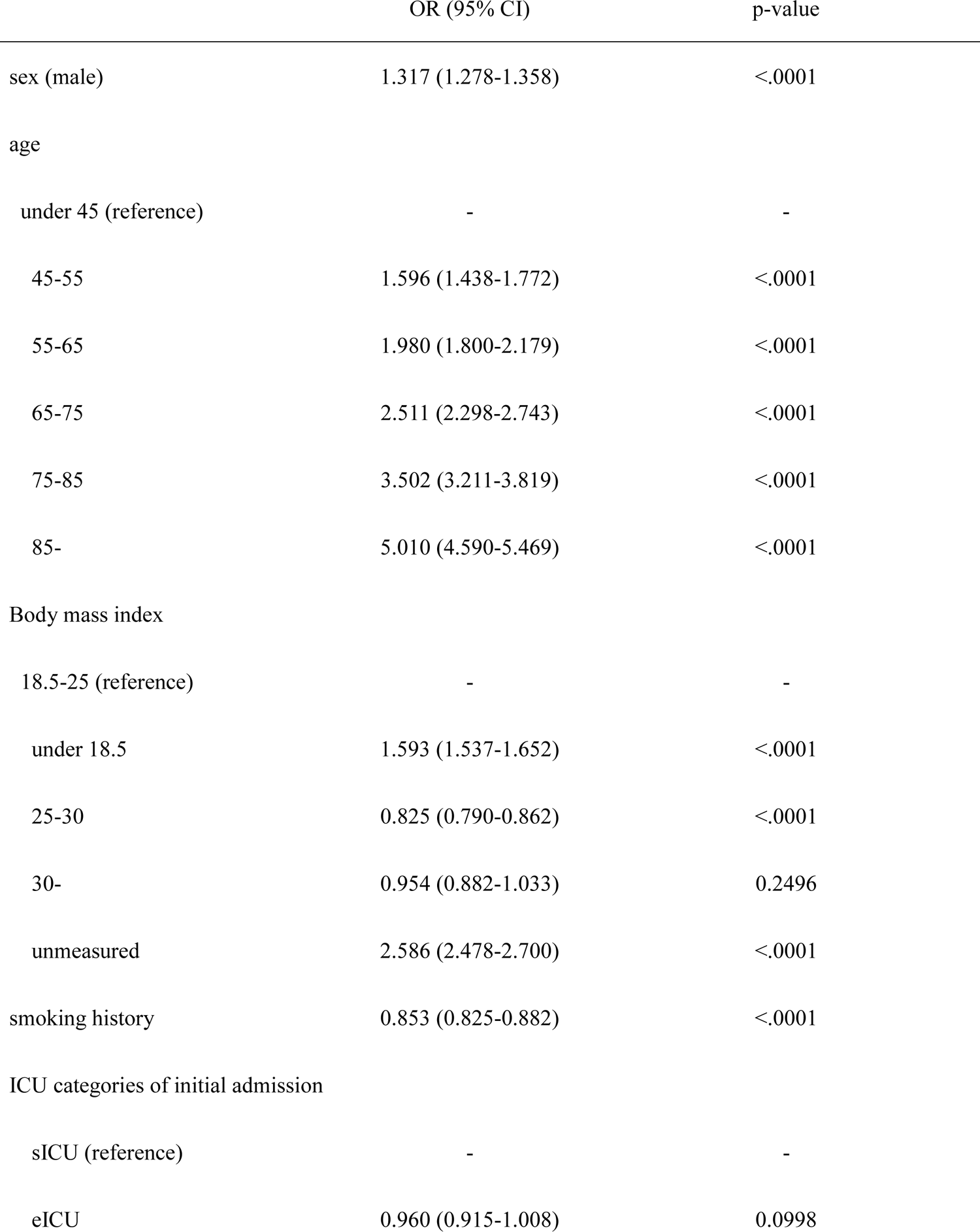
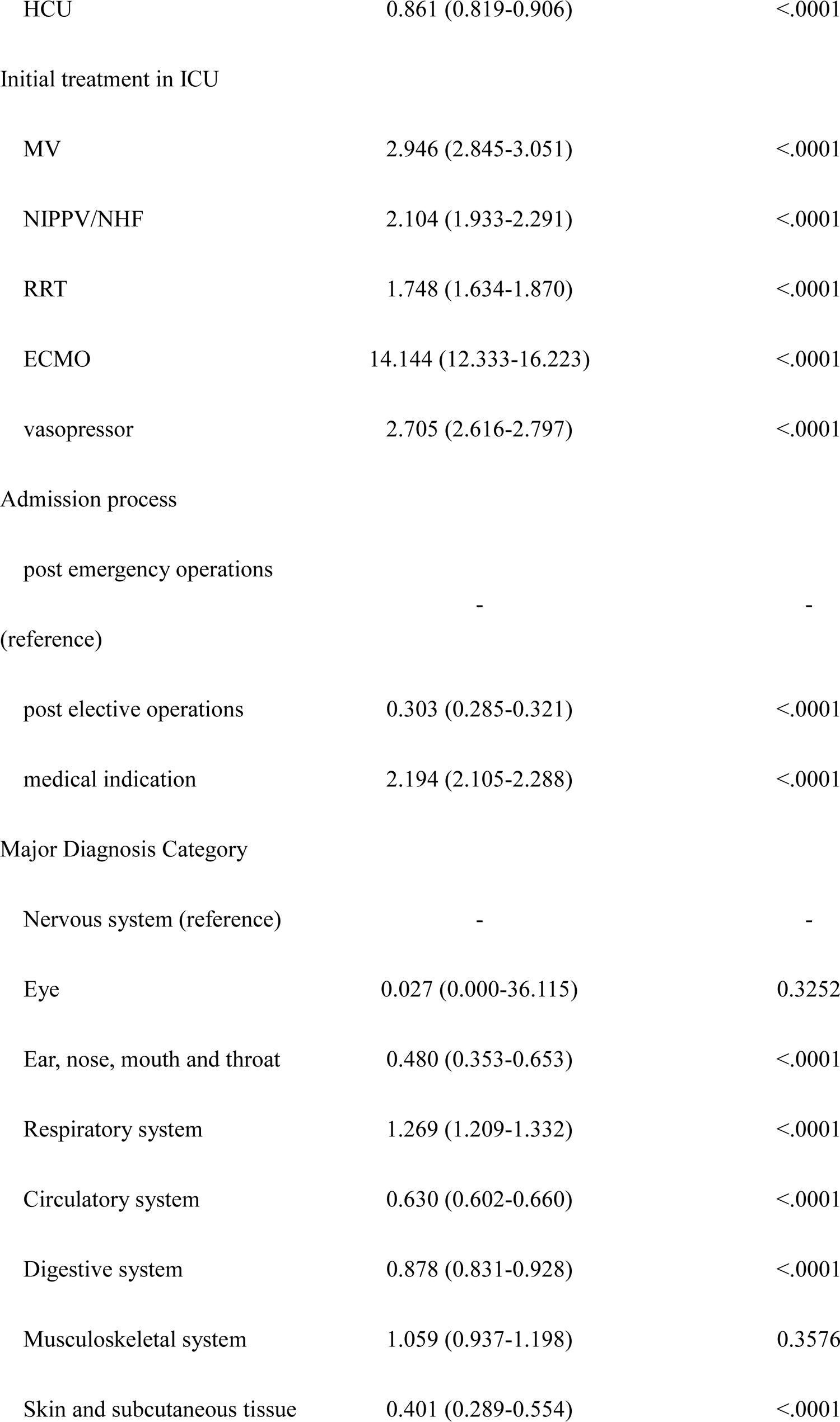
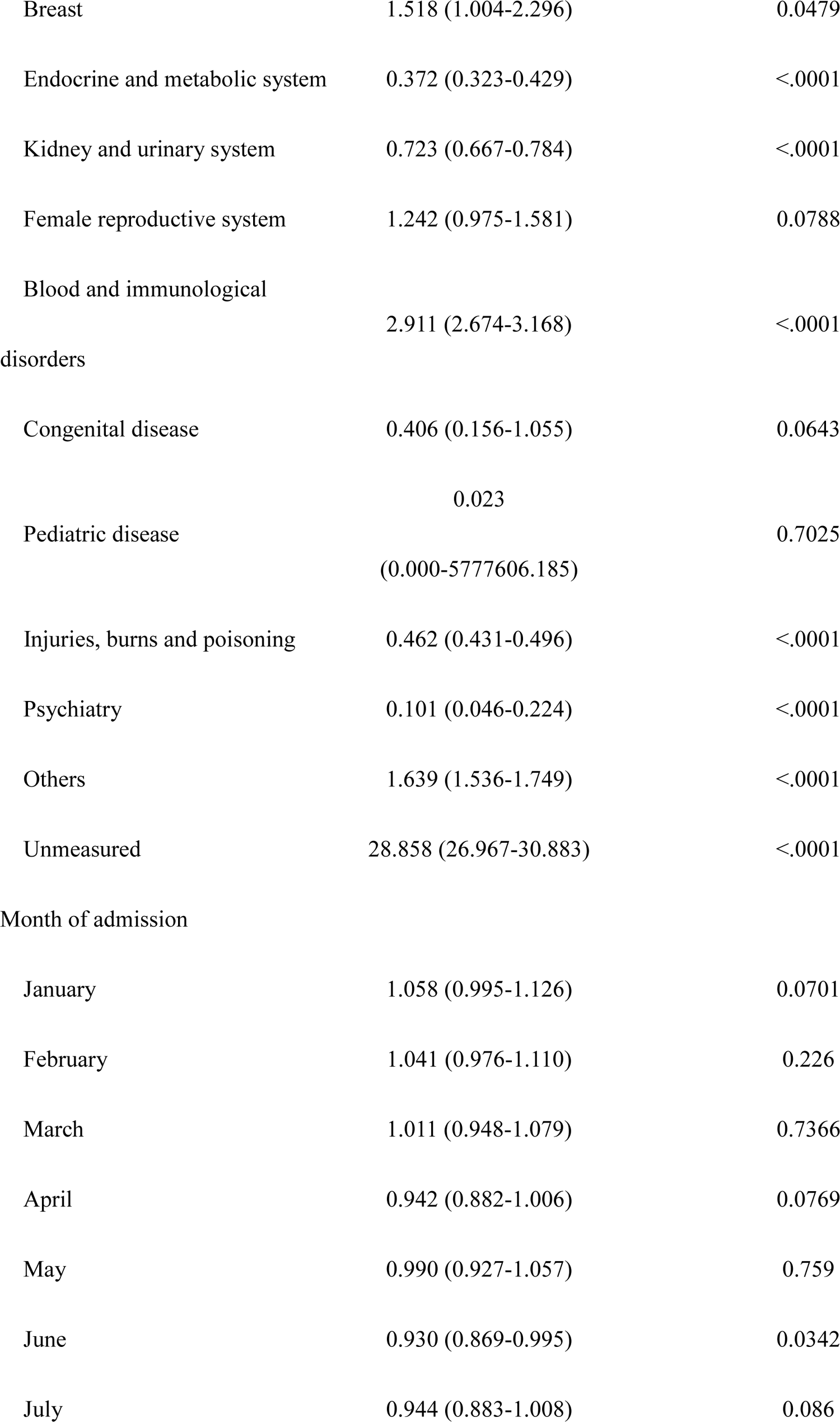
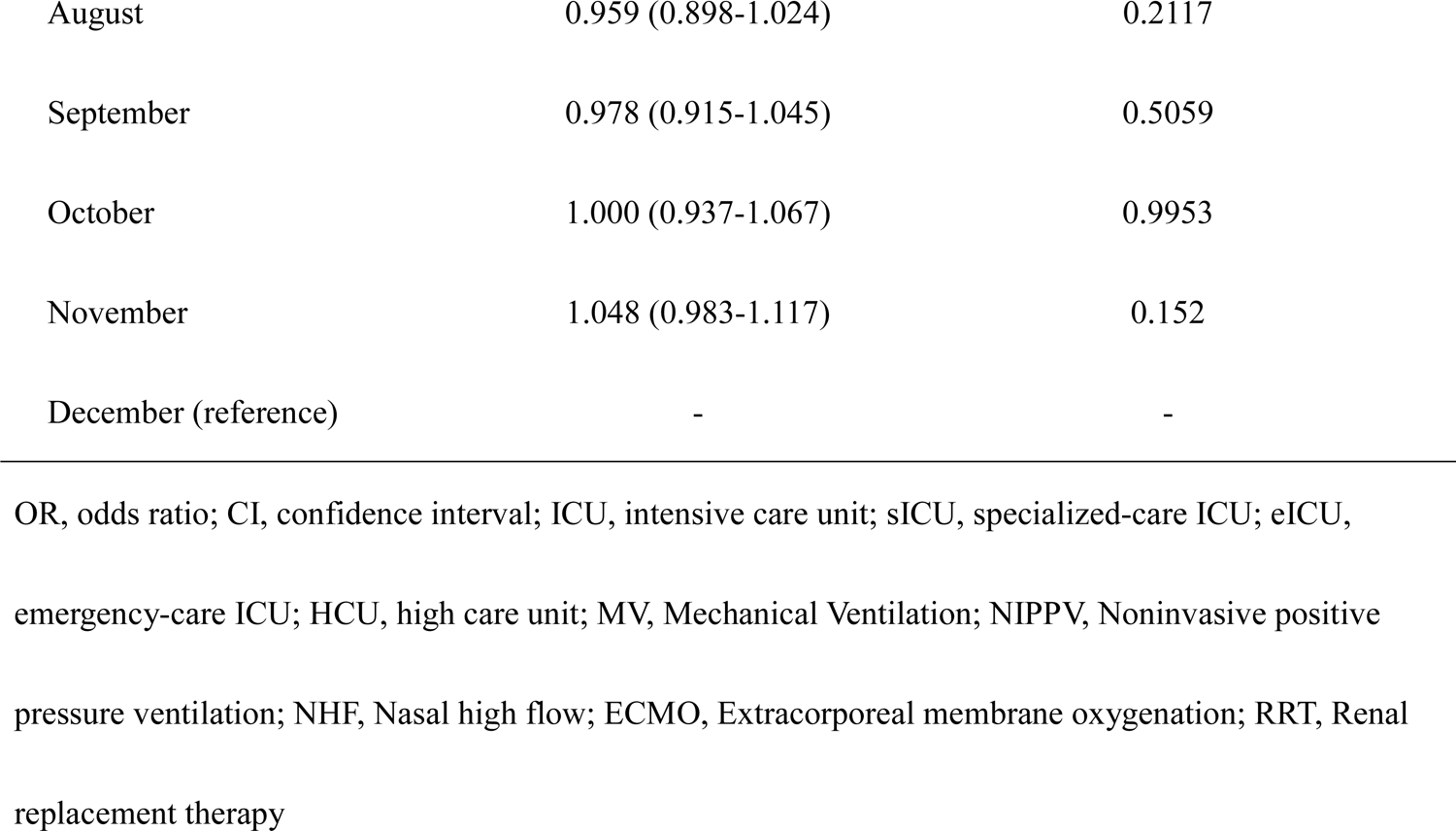
Odds ratios of predictors in the prediction model

**Supplementary Table 6.** Standardized mortality in each wave of the epidemic, stratified by hospital categories

| Time period | Hospital category of acceptance of COVID-19 ICU patients | SMR (95% CI) |
| --- | --- | --- |
| First wave (Apr - Jun 2020) | All hospital | 0.990(0.962-1.019) |
|  | Hospitals, continuously accepting COVID-19 ICU patients | 0.961(0.924-0.999)* |
|  | Hospitals, intermediately accepting COVID-19 ICU patients | 1.018(0.981-1.061) |
|  | Hospitals, accepting few COVID-19 ICU patients | 0.987(0.916-1.059) |
| Second wave (Jul - Sep 2020) | All hospital | 0.979(0.953-1.006) |
|  | Hospitals, continuously accepting COVID-19 ICU patients | 0.975(0.942-1.013) |
|  | Hospitals, intermediately accepting COVID-19 ICU patients | 0.956(0.919-0.996)* |
|  | Hospitals, accepting few COVID-19 ICU patients | 1.057(0.992-1.129) |
| Third wave (Oct 2020 - Mar 2021) | All hospital | 0.996(0.980-1.013) |
|  | Hospitals, continuously accepting COVID-19 ICU patients | 1.001(0.977-1.023) |
|  | Hospitals, intermediately accepting COVID-19 ICU patients | 0.984(0.958-1.009) |
|  | Hospitals, accepting few COVID-19 ICU patients | 1.017(0.977-1.061) |
| fourth wave (Apr 2021 - Jul 2021) | All hospital | 0.989(0.964-1.014) |
|  | Hospitals, continuously accepting COVID-19 ICU patients | 0.963(0.933-0.996)* |
|  | Hospitals, intermediately accepting COVID-19 ICU patients | 1.007(0.972-1.044) |
|  | Hospitals, accepting few COVID-19 ICU patients | 1.011(0.948-1.071) |

SMR, standardized mortality ratio; COVID-19, Coronavirus disease 2019; ICU, intensive care unit
\* indicates statistically significantly different from 1.

## REFERENCES

1. Islam N, Shkolnikov VM, Acosta RJ, et al. Excess deaths associated with covid-19 pandemic in 2020: age and sex disaggregated time series analysis in 29 high income countries. BMJ. 2021;373:n1137.

2. Bontempi E. The europe second wave of COVID-19 infection and the Italy “strange” situation. Environ Res. 2021;193(October 2020):110476.

3. Haldane V, De Foo C, Abdalla SM, et al. Health systems resilience in managing the COVID-19 pandemic: lessons from 28 countries. Nat Med. 2021;27:964–80.

4. Aziz S, Arabi YM, Alhazzani W, et al. Managing ICU surge during the COVID-19 crisis: rapid guidelines. Intensive Care Med. 2020;46(7):1303–25.

5. Novel Coronavirus Response Headquarters (Ver. 2021/7/30). Basic management policy for COVID-19 [Internet]. Tokyo; 2021 [cited 2021 Aug 2]. Available from: https://www.kantei.go.jp/jp/singi/novel_coronavirus/th_siryou/kihon_r_030730.pdf

6. Fadel FA, Al-Jaghbeer M, Kumar S, et al. The impact of the state of Ohio stay-at-home order on non-COVID-19 intensive care unit admissions and outcomes. Anaesthesiol Intensive Ther. 2020;52(3):249–52.

7. Zee-Cheng JE, McCluskey CK, Klein MJ, et al. Changes in Pediatric ICU Utilization and Clinical Trends During the Coronavirus Pandemic. Chest. 2021;160(2):529–37.

8. Bagshaw SM, Zuege DJ, Stelfox HT, et al. Association Between Pandemic Coronavirus Disease 2019 Public Health Measures and Reduction in Critical Care Utilization Across ICUs in Alberta, Canada. Crit Care Med. 2022 Mar 1;50(3):353–362.

9. Hoogendoorn ME, Brinkman S, Bosman RJ, et al. The impact of COVID-19 on nursing workload and planning of nursing staff on the Intensive Care: A prospective descriptive multicenter study. Int J Nurs Stud. 2021 Sep;121:104005.

10. Manzano García G, Ayala Calvo JC. The threat of COVID-19 and its influence on nursing staff burnout. J Adv Nurs. 2021;77(2):832–44.

11. Huespe IA, Marco A, Prado E, et al. Changes in the management and clinical outcomes of critically ill patients without COVID-19 during the pandemic. Rev Bras Ter Intensiva. 2021;33(1):68–74.

12. Bologheanu R, Maleczek M, Laxar D, et al. Outcomes of non-COVID-19 critically ill patients during the COVID-19 pandemic: A retrospective propensity score-matched analysis. Wien Klin Wochenschr. 2021;133:942–50.

13. Wilcox ME, Rowan KM, Harrison DA, et al. Does Unprecedented ICU Capacity Strain, As Experienced During the COVID-19 Pandemic, Impact Patient Outcome? 2022; Publish Ahead of Print:1–9.

14. Gonzenbach TP, McGuinness SP, Parke RL, et al. Impact of Nonpharmaceutical Interventions on ICU Admissions during Lockdown for Coronavirus Disease 2019 in New Zealand - A Retrospective Cohort Study. Crit Care Med. 2021 Oct 1;49(10):1749–1756.

15. Karako K, Song P, Chen Y, et al. Overview of the characteristics of and responses to the three waves of COVID-19 in Japan during 2020-2021. Biosci Trends. 2021;15(1):1–8.

16. Ministry of Health, Labour, and Welfare. Domestic situation of COVID-19 [Internet]. Tokyo; 2021 [cited 2022 Jan 18]. Available from: https://www.mhlw.go.jp/stf/covid-19/open-data.html

17. Cabinet Secretariat. Emergency Declaration [Internet]. [cited 2022 Jan 18]. Available from: https://corona.go.jp/emergency/

18. Marshall JC, Bosco L, Adhikari NK, et al. What is an intensive care unit? A report of the task force of the World Federation of Societies of Intensive and Critical Care Medicine. J Crit Care. 2017;37:270–6.

19. Iwashita Y, Yamashita K, Ikai H, et al. Epidemiology of mechanically ventilated patients treated in ICU and non-ICU settings in Japan: a retrospective database study. Crit Care. 2018;22(1):329.

20. Yamashita K, Ikai H, Nishimura M, et al. Effect of certified training facilities for intensive care specialists on mortality in Japan. Crit Care Resusc. 2013;15(1):28–32.

21. Mohammed MA, Manktelow BN, Hofer TP. Comparison of four methods for deriving hospital standardised mortality ratios from a single hierarchical logistic regression model. Stat Methods Med Res. 2016;25(2):706–15.

22. Morishita T, Takada D, Shin J, et al. Trends, Treatment Approaches, and In-Hospital Mortality for Acute Coronary Syndrome in Japan During the Coronavirus Disease 2019 Pandemic. J Atheroscler Thromb. 2021;28:1–11.

23. Nagano H, Takada D, Shin J, et al. Hospitalization of mild cases of community-acquired pneumonia decreased more than severe cases during the COVID-19 pandemic. Int J Infect Dis. 2021;106(January):323–328.

24. Okuno T, Takada D, ho Shin J, et al. Surgical volume reduction and the announcement of triage during the 1st wave of the COVID-19 pandemic in Japan: a cohort study using an interrupted time series analysis. Surg Today 2021;21:1–8.

25. Headquarter against COVID-19, Ministry of Health, Labour, and Welfare. Management of the in-hosptail health care systems to deal with the expansion of patients infected with the novel Corona virus. (ver.2, March 26. 2021) [Internet]. 2021 [cited 2021 Sep 16]. Available from: https://www.mhlw.go.jp/content/000614595.pdf

26. Solomon MD, Nguyen-Huynh M, Leong TK, et al. Changes in Patterns of Hospital Visits for Acute Myocardial Infarction or Ischemic Stroke During COVID-19 Surges. JAMA. 2021 Jul 6;326(1):82–84.

27. Katsouras C, Tsivgoulis G, Papafaklis M, et al. Persistent decline of hospitalizations for acute stroke and acute coronary syndrome during the second wave of the COVID-19 pandemic in Greece: collateral damage unaffected. Ther Adv Neurol Disord Orig. 2021;14:1–9.

28. Nomura S, Tanoue Y, Yoneoka D, et al. Mobility Patterns in Different Age Groups in Japan during the COVID-19 Pandemic: a Small Area Time Series Analysis through March 2021. J Urban Heal. 2021;98(5):635–41.

29. Feng L, Zhang T, Wang Q, et al. Impact of COVID-19 outbreaks and interventions on influenza in China and the United States. Nat Commun. 2021;12(1): 3249.

30. OECD. The health impact of COVID-19 in regions [Internet]. 2020 [cited 2021 Aug 31]. Available from: https://www.oecd-ilibrary.org/sites/5442b5fc-en/index.html?itemId=/content/component/5442b5fc-en

